# Knowledge, Attitude, and Practice (KAP), and Acceptance and Willingness to Pay (WTP) for Mosquito-Borne Diseases Control through Sterile Mosquito Release in Bangkok, Thailand

**DOI:** 10.1101/2024.01.23.24301641

**Authors:** Kittayapong Pattamaporn, Ninphanomchai Suwannapa, Namon Jalichandra, Sringernyuang Luechai, Sherer Penchan, Meemon Natthani

**Author notes:** **Corresponding author:** Dr. Pattamaporn Kittayapong Center of Excellence for Vectors and Vector-Borne Diseases 2^nd^ Floor, Science Building 2, Faculty of Science Mahidol University at Salaya 999 Phutthamonthon 4 Road Nakhon Pathom 73170, Thailand.

## Abstract

**Background:** Arboviral diseases such as dengue, chikungunya and Zika are public health concerns worldwide. Prevention and control of these diseases still depend on controlling *Aedes aegypti* mosquito vectors. Sterile insect technique (SIT) and incompatible insect technique (IIT) are environmental friendly approaches that show promising impacts. In order to plan an implementation of SIT/IIT technology, the background knowledge, attitude and practice related to these diseases and their mosquito vectors in the targeted communities are needed.

**Methodology/Principal findings:** In this paper, we conducted the questionnaire surveys on general knowledge, attitude and practice (KAP) related to mosquito-borne diseases, mosquito vectors, as well as prevention and control in 400 sampling households in seven communities located in two districts in Bangkok, Thailand. The acceptance and willingness to pay (WTP) for sterile mosquitoes to be used as an alternative vector control approach was also investigated. Our findings indicated that the surveyed participants had high knowledge on dengue (85.25%) and they were more concerned with the severity of dengue (81%) than chikungunya (42.5%) and Zika (37%). Participants with the ages lower than 35 years old (*p* = 0.047) and the incomes higher than 5,000 THB (*p* = 0.016) had more knowledge on mosquito vectors. Moreover, 47% of respondents had positive attitude toward sterile mosquitoes and their application in vector control even though 45.5% of them had never heard about the technology. However, the majority of them were not willing to pay (52%); and if they had to pay, the maximum would be 1-2 THB per sterile mosquito, as most of them expected to receive free service from the government.

**Conclusions/significance:** The baseline information obtained from this questionnaire survey could be used for planning the sterile mosquito release by public health authorities in Bangkok, Thailand where dengue, chikungunya and Zika were still prevalent.

**Author summary:** A questionnaire survey was conducted in seven communities in Bangkok, Thailand to obtain the baseline information on knowledge, attitude and practice (KAP) related to mosquito-borne diseases, i.e., dengue, chikungunya and Zika, including mosquito vectors and how to control them. The questionnaire also asked about the acceptance and willingness to pay (WTP) for sterile mosquitoes used in controlling mosquito populations. Our results showed that, from the total of 400 sampling households, about 85% of participants were familiar with dengue, the mosquito vectors as well as their prevention and control. Furthermore, participants with lower ages and higher incomes had more knowledge on mosquito vectors (*p* < 0.05). Even though the majority of participants showed positive perception about sterile mosquitoes release in terms of the environment, economic, social and quality of life, but more than half of them were not willing to pay for sterile mosquitoes as they would like to receive them free of charge from the government. In the case that they have to pay, the cost that they could afford was 1-2 THB per sterile mosquito. These findings should be useful for public health authorities in planning to apply the sterile mosquito release as an alternative mosquito control approach in Bangkok, Thailand.

## Introduction

Arthropod-borne or arbovirus diseases are public-health concerns in many regions of the world, including Thailand. Important mosquito-borne diseases affecting humans in Thailand include dengue, chikungunya and Zika [Raksakoon & Potiwat, 2021]. However, there are currently no safe and highly effective vaccines available for preventing these diseases [Huang et al., 2023; Costa et al., 2021]. *Aedes* mosquitoes are the most important arbovirus vectors, and *Ae. aegypti* is the major vector of dengue and Zika virus transmission, while *Ae. albopictus* is the major vector of chikungunya virus in Thailand [Phumee et al., 2019; Thavara et al., 2006; Thavara et al., 2009]. Vector control had been the principal method of preventing vector-borne diseases. It involved collaboration with the non-health sector as engagement of the non-health sector increased sustainability of vector control against vector-borne diseases [Wilson et al., 2020].

Regarding community health problems, the knowledge, attitudes, and practices (KAPs) of the populations played a major role in implementation of the control measures [Alobuia et al., 2015]. Understanding and enhancing households’ KAP was crucial for effective community disease control and prevention initiatives [Zhang et al., 2023]. KAP surveys were conducted in many parts of the world in order to understand the local contexts which should be useful for implementation of vector control for dengue, chikungunya, Zika and malaria [Stefopoulou et al., 2021; Omotayo et al., 2021; Kumaran et al., 2018; Jaramillo-Ramírez et al., 2017; Doblecki-Lewis et al., 2016; Mejía et al., 2016; Alobuia et al., 2015]. Some KAP surveys were used to study dengue vaccine acceptance [Shafie et al., 2023]. Furthermore, some vector control implementers applied it before implementation of the new vector control technology [Stefopoulou et al., 2021].

In Thailand, the control of vector-borne diseases, like elsewhere, largely depended on vector management [DVBD, 2019]. However, due to physiological resistance and behavioral avoidance to insecticides of the primary mosquito vectors of human diseases, persistent threats remained and continued to impose a public health burden to vulnerable populations [Chareonviriyaphap et al., 2013]. In our previous work, we had successfully showed a significant reduction of natural populations of *Ae. aegypti* mosquitoes in semi-rural settings in Thailand by an application of a combined Sterile Insect Technique (SIT) and Incompatible Insect Technique (IIT) approach which relied on open field release of sterile males [Kittayapong et al., 2019]. Therefore, we aimed to conduct a KAP survey in order to collect information regarding community knowledge, attitudes, and perceptions about the risk of mosquito exposure and the acceptance of the new SIT/IIT technology from local residents before releasing sterile mosquitoes in a larger scale application in Bangkok, Thailand. Results from this study will be useful for policy makers to consider integrating this new technology together with the routine methods for vector control in Thailand, especially in the Capital City of Bangkok.

## Materials and methods

### Study area

This study was conducted in Chatuchak (13°49′43″N 100°33′35″E) and Bang Khae (13°41′31″N 100°24′26″E) Districts, Bangkok, Central Thailand with the population densities of 4,673.39 and 4,324.57 per sq.km. respectively. Geographically, Bangkok is in the Chao Phraya River delta in Thailand’s central plains. The river meanders through the city in a southward direction, emptying into the Gulf of Thailand approximately 25 km south of the city center. The area is flat and low-lying, with an average elevation of 1.5 m above sea level. Most of the area was originally swampland, which was gradually drained and irrigated for agriculture via the construction of canals. Bangkok was ranked as the top destination city by international visitor arrivals in the Global Destination Cities Index 2023. Besides, there was a lot of people working in Bangkok and they routinely travel to surrounding provinces in the suburb of Bangkok as they live outside the city center making Bangkok an important hotspot for vector-borne diseases for both local and international visitors.

Seven communities, three communities in Chatuchak (KM11, Khlong Prem, and Pahol Yothin 45 communities) and four communities in Bang Khae (Wat Phrom Suwan, Nimmannorade, Yim Prayoon, and Perm Sap communities) Districts (Figure 1), were selected for this study based on dengue incidences from 2008-2022. Retrospective data on dengue was provided by the Health Department, Bangkok Metropolitan Administration. Briefly, Chatuchak District was more in the city center where KM11 community was located about three kilometers away from the famous Chatuchak week-end market, Pahol Yothin 45 community was two kilometers away from Kasetsart University, and Klong Prem community was only few kilometers away from the Klong Prem central prison. However, for Bang Khae District, it was in the southwest of Bangkok and more rural areas such agricultural plantation, was observed. In terms of household structure, common construction either single or two-story houses built by woods or concrete or sometimes connected building blocks were found. Regarding population, people living in communities in Chatuchak District, were more of those who moved from other parts of the country or other part of Bangkok and lived there for job opportunity, whereas for those who lived in communities in Bang Khae District were more of the local people who stayed or lived there with their family for generations.

### Data collection

In this study, data was collected from April 2022 to August 2023 by using a constructed questionnaire. The questionnaire for the KAP survey and the willingness to pay (WTP) were adopted from instruments developed by the WHO [WHO, 2008], and Yeo and Shafie [Yeo & Shafie, 2018] respectively. For data collection, the research team worked in close contact with the local health volunteers in each community and the health volunteers were in charge of distribution of the questionnaire to the householders based on the availability of householders. The head of each household or an adult household member was requested to fill out and answered all questions by themselves. Also, they were asked to provide a written consent on the information sheet, and the consent form before returning it back to the health volunteers. The completed questionnaires, together with the information sheets and the consent forms from each household within the same community, were collected by the health volunteers and sent by post to the research team for data entry and further analysis.

### Study instrument

A constructed questionnaire was used as a research instrument for this study. It consisted of 126 multiple choice questions and one open question. The questionnaire was mainly divided into five parts. The first part consisted of 10 questions on demographic information, such as age, gender, educational level, marital status, employment status or occupation, monthly income, number of family members, and type and characteristic of house. The second part included 29 questions on awareness and knowledge of dengue, chikungunya, and Zika diseases. It also included questions on sources of information about these diseases as well as disease transmission, disease vectors, mosquito breeding sites, experience of disease infection, management and control of diseases, attitudes towards dengue, chikungunya, and Zika prevention. The third part consisted of 35 questions on knowledge of the SIT/IIT approach for vector control such as source of information about sterile mosquitoes, how to produce sterile mosquitoes, differentiation between sterile and wild mosquitoes, benefit of application of sterile mosquitoes for vector control, frequency for release of the sterile mosquitoes in the fields. In this section, questions also included attitudes towards application of sterile mosquitoes for vector control and factors that people might take into consideration when release sterile mosquitoes in their households or nearby. In addition, 18 questions on the willingness to pay (WTP) were also included in this section to assess willingness-to-pay/demand of the local residents [Biadgilign et al., 2015] for vector control through an application of sterile mosquito release. WTP is the maximum a person or a household would be willing to pay for a good or service [Onwujekwe et al., 2004]. To assess WTP, an artificial market was used to measure consumer preferences by directly asking their willingness to pay or willingness to accept for change in the level of good or service [Brouwer & Bateman, 2005]. The total value of the good, both use and non-use values, and its flexibility facilitate valuation of a wide range of non-market goods could also be captured by this method [Biadgilign et al., 2015]. In this study, a direct open-end format was used to describe for uncovering value by asking respondent about the maximum price that individual respondent is willing to pay for a given treatment of sterile mosquito release. The forth part consisted of 26 questions on impacts of an application of the release of sterile mosquitoes on environment, economy, society and quality of life based on the attitudes and perceptions of the respondents, such as reduction of vector control costs or insecticide applications, feeling happy and safe from disease transmission, etc. Lastly, the fifth part included 25 questions on community engagement and acceptance of the research project such as community participation, willingness of respondents to participate in the research project, duration of the project, problems or difficulties encountered during the project, and level of acceptance and satisfaction to the project. The last two parts, respondents were asked to score from ‘totally agree’ to ‘totally disagree’ as a highest and lowest satisfaction, and ‘do not know’ was also provided for those who had no response for each specific question.

### Ethical consideration

The proposal and all relevant documents in this study were reviewed and approved by the Bangkok Metropolitan Administration Human Research Ethics Committee (BMAHREC E003q/63_EXP). Supporting letters were obtained from the District Health Offices after explaining the purpose and significance of the study. Written consents were also obtained from individual respondents from each participating household.

### Statistical analysis

Data was entered and cleaned using Microsoft Office Excel 2016 and statistical analysis was performed further using SPSS 18.0 (Mahidol University License (Chicago, SPSS Inc.). Descriptive summaries (frequencies and proportions) were calculated. Results were presented as Odds ratio and 95% confidence interval (CI) were used to examine the strength of association with the main variable of interests. Chi-square tests as well as univariate followed by multivariate logistic regressions were performed in order to determine the factors influencing knowledge with a statistical significance of 5%, and the *p*-values of less or equal to 0.05 were considered significant.

## Results

### Participant demographics

Out of 400 participants, higher numbers of women (59.50%) read and fill out the questionnaire at home when compared to men who were usually busy working outside (Table 1). The ages of participants ranged from 18 to 87 years with the mean age of 51 years old. The majority of participants were married (43.50%), and they were head of the family (38.50%) with the highest level of education at the primary school (27.50%). The primary occupation of people surveyed was government or state enterprise officer (25.00%), followed by laborer (23.75%), because this study was conducted in a community supervised by the State Railway of Thailand, the state enterprise under the Ministry of Transportation. Most participants earned monthly income of 5,001-10,000 THB (US$ 142 – 285) and they lived in their own houses (36.00%) followed by state enterprise residences (35.75%).

**Table 1.** Demographic information of surveyed participants living in Bangkok, Thailand.

| **Characteristics** | **% (N = 400)** | **Median (range)** |
| --- | --- | --- |
| **Gender** |  |  |
| Female | 59.50 (238) | - |
| Male | 40.50 (162) | - |
| **Age group (years)** |  |  |
| 18-34 | 13.75 (55) | 27 (18-34) |
| 35-54 | 35.25 (141) | 46 (35-54) |
| > 55 | 38.5 (154) | 62 (55-87) |
| Unknown / Not answer | 12.50 (50) | - |
| **Marital status** |  |  |
| Single | 24.50 (98) | - |
| Boyfriend / girlfriend | 10.50 (42) | - |
| Married | 43.50 (174) | - |
| Divorce | 3.00 (12) | - |
| Widow | 7.75 (31) | - |
| Unknown / Not answer | 10.75 (43) | - |
| **Family status** |  |  |
| Head of family | 38.50 (154) | - |
| Spouse | 28.25 (113) | - |
| Son/daughter | 9.25 (37) | - |
| Relatives | 4.50 (18) | - |
| Others | 4.00 (16) | - |
| Unknown/ Not answer | 15.50 (62) | - |
| **Education** |  |  |
| None | 3.25 (13) | - |
| Primary school | 27.50 (110) | - |
| Secondary school | 20.50 (82) | - |
| High school | 14.50 (58) | - |
| Bachelor's degree | 20.75 (83) | - |
| Higher than Bachelor's degree | 4.00 (16) | - |
| Unknown/ Not answer | 9.50 (38) | - |
| **Occupation** |  |  |
| Laborer | 23.75 (95) | - |
| Agriculture/farmer | 0.25 (1) | - |
| Trading/merchant | 14.00 (56) | - |
| Government/State Enterprise officer | 25.00 (100) | - |
| Company employee | 3.75 (15) | - |
| Housekeeper | 13.50 (54) | - |
| Others | 8.5 (34) | - |
| Unknown/ Not answer | 11.25 (45) | - |
| **Monthly income (THB) (US\$ 1 = 34 THB)** | | |
| < 5,000 | 8.50 (34) | - |
| 5,001-10,000 | 23.00 (92) | - |
| 10,001-15,000 | 15.00 (60) | - |
| 15,001-20,000 | 13.25 (53) | - |
| 20,001-30,000 | 6.50 (26) | - |
| > 30,000 | 10.00 (40) | - |
| Unknown/ Not answer | 23.75 (71) | - |
| **Number of family members** |  |  |
| 1 - 2 | 17.25 (69) | 2 (1-2) |
| 3 – 4 | 37.25 (149) | 4 (3-4) |
| > 5 | 27.75 (111) | 5 (5-12) |
| Unknown/ Not answer | 17.75 (71) | - |
| **Characteristic of house** |  |  |
| Rental house | 17.25 (69) | - |
| Government/State enterprise residence | 35.75 (143) | - |
| Own house | 36.00 (144) | - |
| Unknown/ Not answer | 11.00 (44) | - |
| **Type of house** |  |  |
| House with garden | 27.50 (110) | - |
| One-story house with high basement | 9.75 (39) | - |
| Two-story or more house | 9.75 (39) | - |
| Commercial building | 1.50 (6) | - |
| Apartment/Dormitory | 1.75 (7) | - |
| Row house | 14.00 (56) | - |
| Others | 7.00 (28) | - |
| Unknown/ Not answer | 28.75 (115) | - |

### Knowledge on dengue, chikungunya, and Zika

Amongst the surveyed participants, they were most familiar with dengue (85.25%) but were less familiar with chikungunya (39.75%) and Zika (33.75%) (Table 2). Participants received information about these diseases mainly from television (20-25%), followed by health officials or village health volunteers (18-20%). The majority of them were most concerned about the severity of dengue (81.00%), but much less was concerned about chikungunya (42.50%), and Zika (37.00%).

**Table 2.** Knowledge on dengue, chikungunya and Zika of the surveyed participants living in Bangkok, Thailand.

| **Characteristics** |  |  |  |
| --- | --- | --- | --- |
|  | **Dengue** | **Chikungunya** | **Zika** |
| **Have you heard about these diseases?** | **% (N = 400)** | **% (N = 400)** | **% (N = 400)** |
| Yes | 85.25 (341) | 39.75 (159) | 33.75 (135) |
| Never | 6.50 (26) | 35.00 (140) | 39.75 (159) |
| Unknown/Not answer | 8.25 (33) | 25.25 (101) | 26.50 (106) |
| **What are your sources of information about these diseases?** |  |  |  |
| Radio | 11.25 (45) | 5.25 (21) | 4.00 (16) |
| Television | 45.00 (180) | 23.75 (95) | 20.25 (81) |
| Newspapers | 3.25 (13) | 0.75 (3) | 0.75 (3) |
| Online Media | 9.25 (37) | 6.50 (26) | 6.75 (27) |
| Poster / Brochure | 0.75 (3) | 0 (0) | 0.25 (1) |
| Health official/Village health volunteer | 20.75 (83) | 19.00 (76) | 18.00 (72) |
| Neighbor / Acquaintance | 2.00 (8) | 2.50 (10) | 1.25 (5) |
| Workplace / School | 1.50 (6) | 0.50 (2) | 0.25 (1) |
| Unknown / Not answer | 6.25 (25) | 41.75 (167) | 48.50 (194) |
| **Do you think these diseases are severe?** |  |  |  |
| Severe | 81.00 (324) | 42.50 (170) | 37.00 (148) |
| Mild | 4.50 (18) | 8.25 (33) | 6.25 (25) |
| Unknown/Not answer | 14.50 (58) | 49.25 (197) | 56.75 (227) |
| **What kinds of mosquitoes carry these diseases?** |  |  |  |
| *Aedes* mosquitoes | 87.25 (349) | 42.25 (169) | 35.50 (142) |
| *Culex* mosquitoes | 0.75 (3) | 0.50 (2) | 1.25 (5) |
| *Anopheles* mosquitoes | 0.50 (2) | 4.75 (19) | 1.00 (4) |
| All kinds of mosquitoes | 4.25 (17) | 7.00 (28) | 10.25 (41) |
| Unknown/Not answer | 7.25 (29) | 45.50 (182) | 52.00 (208) |
| **Identify breeding sites of *Aedes* mosquitoes** |  |  |  |
| Stagnant water | 78.50 (314) | 43.25 (173) | 39.25 (157) |
| Flowing tides | 1.50 (6) | 1.50 (6) | 0.73 (3) |
| Dirty places | 2.25 (9) | 3.50 (14) | 3.50 (14) |
| Water containers | 11.25 (45) | 6.00 (24) | 5.25 (21) |
| Wasteland | 0.25 (1) | 1.25 (5) | 1.00 (4) |
| Garbage disposal areas | 0.50 (2) | 0.75 (3) | 0.50 (2) |
| Drainage pipe system | 1.50 (6) | 2.00 (8) | 2.00 (8) |
| Unknown/Not answer | 4.25 (17) | 41.75 (167) | 47.75 (191) |
| **How are these diseases transmitted from person to person?** |  |  |  |
| By mosquito bites | 81.00 (324) | 46.75 (187) | 40.50 (162) |
| By touching each other | 3.00 (12) | 2.25 (9) | 2.25 (9) |
| By water | 2.00 (8) | 1.50 (6) | 1.00 (4) |
| By flies | 0.50 (2) | 0.50 (2) | 0.75 (3) |
| By other animals | 2.00 (8) | 1.25 (5) | 2.00 (8) |
| Unknown/Not answer | 11.50 (46) | 47.75 (191) | 53.50 (214) |
| **When do you think *Aedes* mosquitoes bite people?** |  |  |  |
| Day | 64.00 (265) | 36.00 (144) | 31.00 (124) |
| Night | 5.25 (21) | 4.00 (16) | 3.50 (14) |
| Both day and night | 20.00 (80) | 13.00 (52) | 11.50 (46) |
| Evening / Dusk | 3.75 (15) | 3.00 (12) | 2.75 (11) |
| Unknown / Not answer | 7.00 (28) | 44.00 (176) | 51.25 (205) |
| **Have you ever suffered from these diseases?** |  |  |  |
| Yes | 18.75 (75) | 2.25 (9) | 1.00 (4) |
| No | 71.00 (284) | 64.75 (259) | 63.00 (252) |
| Unknown/Not answer | 10.25 (41) | 33.00 (132) | 36.00 (144) |
| **Do you have any acquaintances who have suffered from these diseases?** |  |  |  |
| Yes | 48.25 (193) | 14.00 (56) | 6.00 (24) |
| No | 41.00 (164) | 47.75 (191) | 51.50 (206) |
| Unknown / Not answer | 10.75 (43) | 38.25 (153) | 42.50 (170) |

Knowledge about disease vectors, transmission and breeding sites was quite high. The vast majority of people were able to identify *Aedes* mosquitoes as the major vector of dengue (87.25%), but much less was able to identify chikungunya (42.25%) and Zika (35.50%) mosquito vectors. However, when asked about chikungunya (45.50%) and Zika (52.00%), many people had more doubt or they preferred not to answer, especially for Zika (Table 2). The majority of people were able to identify the stagnant water or water containers as the breeding sites of *Aedes* mosquitoes. They were able to identify correctly that the diseases were transmitted through mosquito bites and they could notice that *Aedes* mosquitoes bit during the daytime (dengue = 64.00%, chikungunya = 36.00%, Zika = 31.00%). However, some of participants still believed that mosquito vectors bit at night (dengue = 5.25%, chikungunya = 4.00%, Zika = 3.50%) or both day and night (dengue = 20.00%, chikungunya = 13.00%, Zika = 11.50%). When asked about experience of getting infected with these arboviruses, the majority of participants had not yet been infected with dengue (71.00%), chikungunya (64.50%) and Zika (63.00%).

### Prevention and control measures for dengue, chikungunya, and Zika

When asked about getting rid of breeding sites in or around their houses, the majority of respondents put the lids on tightly for all water containers (11.25%), followed by disposing discarded water containers and garbage (8.65%), and changing water in the water container weekly (5.85%) (Table 3). When asked about the methods of prevention from mosquito bites, the majority of respondents chose to sleep under the bed nets (9.50%) followed by installing mosquito screens (8.55%), using mosquito repellent coils (7.55%), and turning on the fan to prevent mosquito bites (7.35%).

**Table 3.** Prevention and control measures for dengue, chikungunya, and Zika of the surveyed participants living in Bangkok, Thailand.

| Characteristics | % (N = 400) |
| --- | --- |
| **How do you get rid of the breeding sites for mosquito larvae in and around your homes?** |  |
| Put lids on all water containers tightly | 11.25 (45) |
| Change water in the water containers weekly | 5.85 (23) |
| Discarded water container disposal/ garbage disposal | 8.65 (35) |
| Brush and scrub inside the water containers | 1.85 (7) |
| Release fish to consume larvae in water containers | 2.75 (11) |
| Put larvicides/chemicals in water containers | 2.75 (11) |
| Scoop mosquito larvae out of water containers | 1.10 (4) |
| Did not do anything | 1.10 (4) |
| Unknown | 0.65 (3) |
| Not answer | 64.15 (257) |
| **What methods do you use to protect yourself and your family members from mosquito bites?** |  |
| Sleep under mosquito nets | 9.50 (38) |
| Use mosquito repellent coils | 7.55 (30) |
| Install mosquito screens | 8.55 (34) |
| Turn on the fan to prevent mosquito bites | 7.35 (29) |
| Use a mosquito shock machine/ mosquito mat | 3.75 (15) |
| Use insecticide-impregnated mosquito nets | 0.70 (3) |
| Apply mosquito repellent lotion when entering the forest/ garden | 4.25 (17) |
| Wear long-sleeved shirts and long pants when entering the forest/garden | 1.50 (6) |
| Did not do anything | 0.25 (1) |
| Others | 0.35 (1) |
| Unknown | 0.75 (3) |
| Not answer | 55.50 (222) |

### Attitude towards dengue, chikungunya, and Zika

The surveyed participants were almost all agreed or totally agreed to keep the house clean (91.00%), and to empty and scrub the water storage containers once a week (89.00%) (Table 4). However, they were all agreed or totally agreed that elimination of the mosquito breeding sites was difficult (67.50%). In case of having dengue, chikungunya, and Zika patients in the houses, the respondents believed that they had to cooperate in order to get rid of the mosquito breeding sites (86.75%). People also believed that health officials play an important role in preventing dengue, chikungunya, and Zika infections in the communities (82.25%). Some respondents believed that disposal of mosquito breeding sites was the sole responsibility of the health officials (35.75%) while others believed the opposite (40.00%). However, some respondents were uncertain (19.25%). The vast majority of respondents believed that sleeping under mosquito nets could prevent them from mosquito bites (63.75%). People believed that it could be life-threatening (84.00%) when getting infected with dengue, chikungunya, or Zika viruses without prompt treatment; and the best prevention method was to prevent from mosquito bites (78.50%).

**Table 4.** Attitude towards dengue, chikungunya, and Zika of the surveyed participants living in Bangkok, Thailand.

| Characteristics | % (N = 400) |
| --- | --- |
| **It is essential to keep the house and surrounding areas clean.** |  |
| Totally agree | 61.75 (247) |
| Agree | 29.25 (117) |
| Uncertain | 4.50 (18) |
| Disagree | 0 (0) |
| Strongly disagree | 0 (0) |
| Unknown/Not answer | 4.50 (18) |
| **It is the right thing to do to empty and scrub water storage containers once a week.** |  |
| Totally agree | 53.00 (121) |
| Agree | 36.00 (144) |
| Uncertain | 6.75 (27) |
| Disagree | 0 (0) |
| Strongly disagree | 0 (0) |
| Unknown/Not answer | 4.25 (17) |
| **It is difficult to eliminate breeding sites of the mosquito vectors of dengue, chikungunya and Zika viruses.** |  |
| Totally agree | 36.25 (145) |
| Agree | 31.25 (125) |
| Uncertain | 18.25 (73) |
| Disagree | 7.75 (31) |
| Strongly disagree | 1.25 (5) |
| Unknown/Not answer | 5.25 (21) |
| **Households with dengue, chikungunya and Zika patients must cooperate to eliminate mosquito breeding sites** |  |
| Totally agree | 45.25 (181) |
| Agree | 41.50 (166) |
| Uncertain | 7.25 (29) |
| Disagree | 1.25 (5) |
| Strongly disagree | 0.25 (1) |
| Unknown/Not answer | 4.50 (18) |
| **Health officials play a critical role in preventing dengue, chikungunya, and Zika at the community level** |  |
| Totally agree | 46.50 (186) |
| Agree | 35.75 (143) |
| Uncertain | 9.50 (38) |
| Disagree | 3.25 (13) |
| Strongly disagree | 0.50 (2) |
| Unknown/Not answer | 4.50 (18) |
| **Disposal of mosquito breeding sites is the sole responsibility of health officials** |  |
| Totally agree | 20.75 (83) |
| Agree | 15.00 (60) |
| Uncertain | 19.25 (77) |
| Disagree | 28.75 (115) |
| Strongly disagree | 11.25 (45) |
| Unknown/Not answer | 5.00 (20) |
| **Sleeping under mosquito nets can prevent dengue, chikungunya and Zika viruses** |  |
| Totally agree | 26.50 (106) |
| Agree | 37.25 (149) |
| Uncertain | 25.50 (102) |
| Disagree | 5.25 (21) |
| Strongly disagree | 1.00 (4) |
| Unknown/Not answer | 4.50 (18) |
| **It could be life-threatening if you are sick with dengue or chikungunya or Zika and do not treated quickly** |  |
| Totally agree | 45.25 (181) |
| Agree | 38.75 (155) |
| Uncertain | 10.50 (42) |
| Disagree | 0.50 (2) |
| Strongly disagree | 0.75 (3) |
| Unknown/Not answer | 4.25 (17) |
| **The best method to prevent dengue, chikungunya and Zika is to avoid mosquito bites** |  |
| Totally agree | 42.00 (168) |
| Agree | 36.50 (146) |
| Uncertain | 15.25 (61) |
| Disagree | 1.00 (4) |
| Strongly disagree | 0 (0) |
| Unknown/Not answer | 5.25 (21) |

### Practice of vector control measures

The vast majority of participants always or occasionally practiced prevention and control measures at home. The majority of them experienced having mosquito larvae in water storage containers (80.00%) (Table 5). When finding larvae, some people always (40.50%) or occasionally (28.75%) removed larvae from the containers while others did nothing (19.50%). People always (31.25%) or occasionally (25.00%) added abate sand in water containers when they find larvae inside, while many people still did nothing (32.00%). Many people always (55.00%) or occasionally (29.50%) cleaned water containers when they found larvae. Many people always (43.00%) or occasionally (35.75%) changed water or cleaned water containers weekly. People occasionally (37.25%) or always (22.00%) changed water or added vinegar or detergent or salt in pantry leg saucers every week to get rid of mosquito larvae. When finding mosquito breeding sites, i.e., garbage, coconut shells, cans, tires, people occasionally (39.50%) or always (29.50%) turned them upside down in order to get rid of the breeding sites. People applied guppy fishes in water containers (46.75%), however, many of them did nothing (44.00%). Many of them never covered water containers with screen (47.75%) while some covered them with screen (40.50%). The majority of them slept under mosquito nets (72.25%), while others never did (22.25%). Lastly, the majority of them used mosquito repellent coils (72.25%) while others never did (22.25%).

**Table 5.** Practice in vector control by the surveyed participants living in Bangkok, Thailand.

| Characteristics | % (N = 400) |
| --- | --- |
| **Have you ever explored mosquito larvae in drinking water containers, cement basins in bathrooms / toilets, or other water storage containers?** |  |
| Always | 39.25 (157) |
| Occasionally | 40.75 (163) |
| Never did | 14.00 (56) |
| Not eligible | 1.50 (6) |
| Unknown/Not answer | 4.50 (18) |
| **If you find larvae in drinking water containers, cement basins in bathrooms / toilets or other water storage containers, have you ever:** |  |
| **(1) Remove larvae** |  |
| Always | 40.50 (162) |
| Occasionally | 28.75 (115) |
| Never did | 19.50 (78) |
| Not eligible | 2.00 (8) |
| Unknown/Not answer | 9.25 (37) |
| **(2) Add abate sand** |  |
| Always | 31.25 (125) |
| Occasionally | 25.00 (100) |
| Never did | 32.00 (128) |
| Not eligible | 3.25 (13) |
| Unknown/Not answer | 8.50 (34) |
| **(3) Clean water containers** |  |
| Always | 55.00 (220) |
| Occasionally | 29.50 (118) |
| Never did | 8.00 (32) |
| Not eligible | 2.25 (9) |
| Unknown/Not answer | 5.25 (21) |
| **Do you change water and wash flower vases, spotted betel vases, plant pot saucers weekly?** |  |
| Always | 43.00 (172) |
| Occasionally | 35.75 (143) |
| Never did | 10.00 (40) |
| Not eligible | 6.75 (27) |
| Unknown/Not answer | 4.50 (18) |
| **Do you change water or add vinegar or detergent or salt in pantry leg saucers weekly?** |  |
| Always | 22.00 (88) |
| Occasionally | 37.25 (149) |
| Never did | 31.25 (125) |
| Not eligible | 5.00 (20) |
| Unknown/Not answer | 4.50 (18) |
| **Have you ever surveyed water-holding wastes such as coconut shells, cans, tires, in your household area, and have you overturned, burned, landfilled, or destroyed them weekly?** |  |
| Always | 29.50 (118) |
| Occasionally | 39.50 (158) |
| Never did | 22.25 (89) |
| Not eligible | 4.00 (16) |
| Unknown/Not answer | 4.75 (19) |
| **Do you put guppy fishes in water containers such as jars or cement basins?** |  |
| Always | 20.00 (80) |
| Occasionally | 26.75 (107) |
| Never did | 44.00 (176) |
| Not eligible | 4.00 (16) |
| Unknown/Not answer | 5.25 (21) |
| **Do you use lids to cover all jars that are filled with water?** |  |
| Always | 16.50 (66) |
| Occasionally | 24.00 (96) |
| Never did | 47.75 (191) |
| Not eligible | 6.25 (25) |
| Unknown/Not answer | 5.50 (22) |
| **Do you sleep under a mosquito net at night?** |  |
| Always | 42.25 (169) |
| Occasionally | 26.00 (104) |
| Never did | 22.25 (89) |
| Not eligible | 5.00 (20) |
| Unknown/Not answer | 4.50 (18) |
| **Do you use mosquito repellent coils to repel mosquitoes?** |  |
| Always | 42.25 (169) |
| Occasionally | 26.00 (104) |
| Never did | 22.25 (89) |
| Not eligible | 5.00 (20) |
| Unknown/Not answer | 4.50 (18) |

### Knowledge on sterile mosquitoes

The vast majority of people never heard about sterile mosquitoes or related issues (45.50%) but some of them had received information about it (37.75%) (Table 6). When asked those who heard about sterile mosquitoes, the major source of information was from municipal officers (19.90%), followed by leaflets or flyers (16.75%), village broadcast towers (13.61%) and local newspapers (12.57%). For those who heard about sterile mosquitoes, they most likely shared it to their acquaintances (58.50%). However, some of them were still uncertain (14.50%) to share it.

**Table 6.**
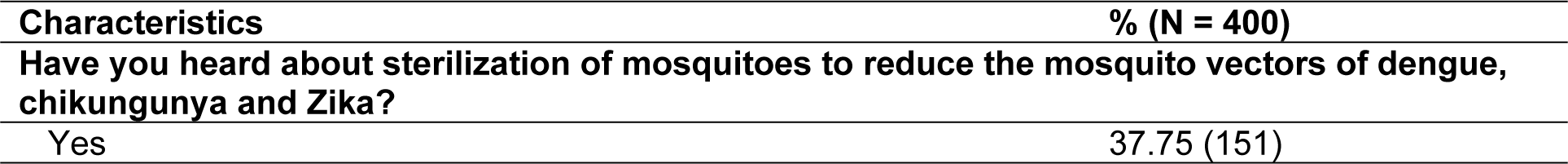
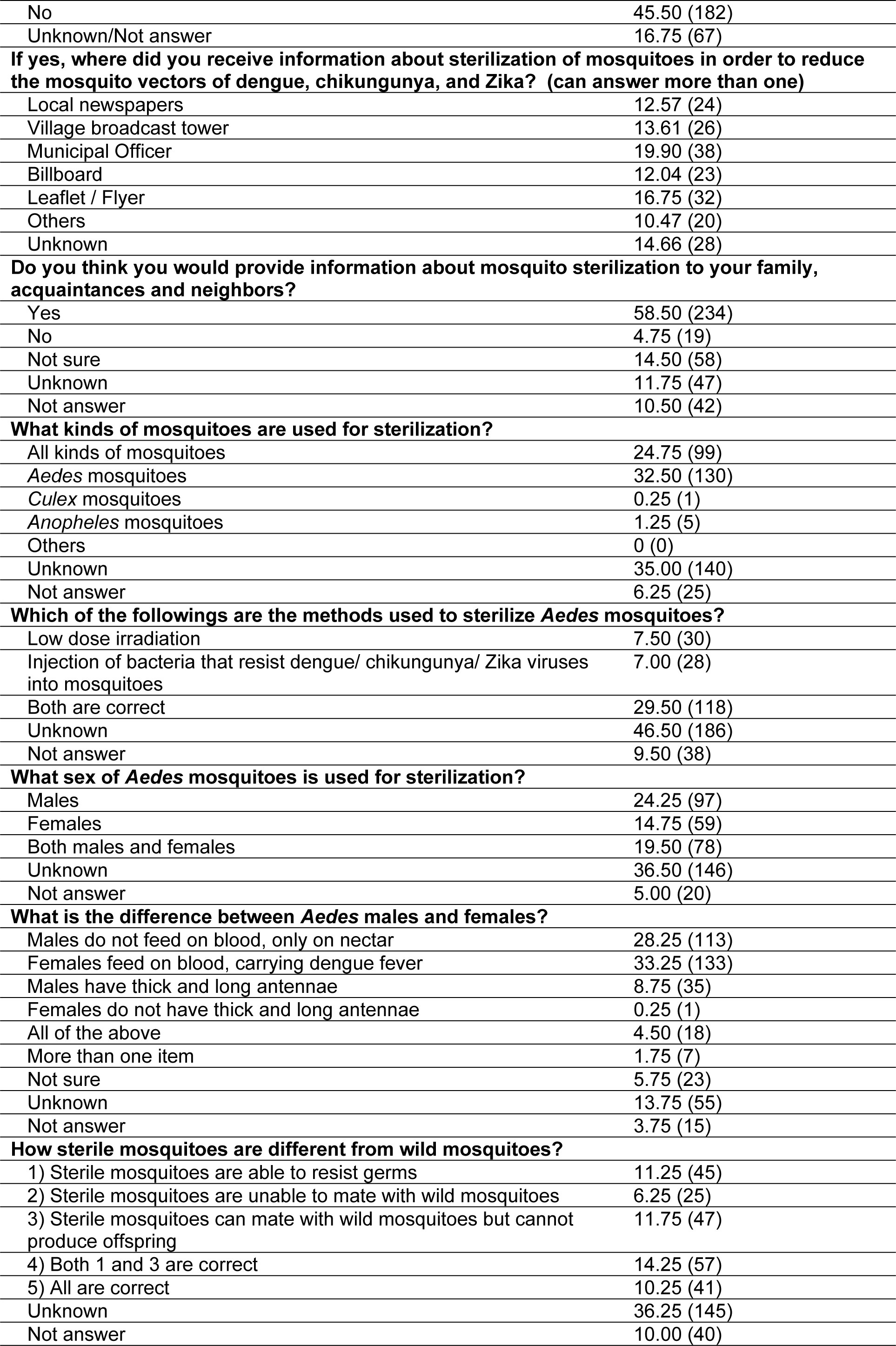
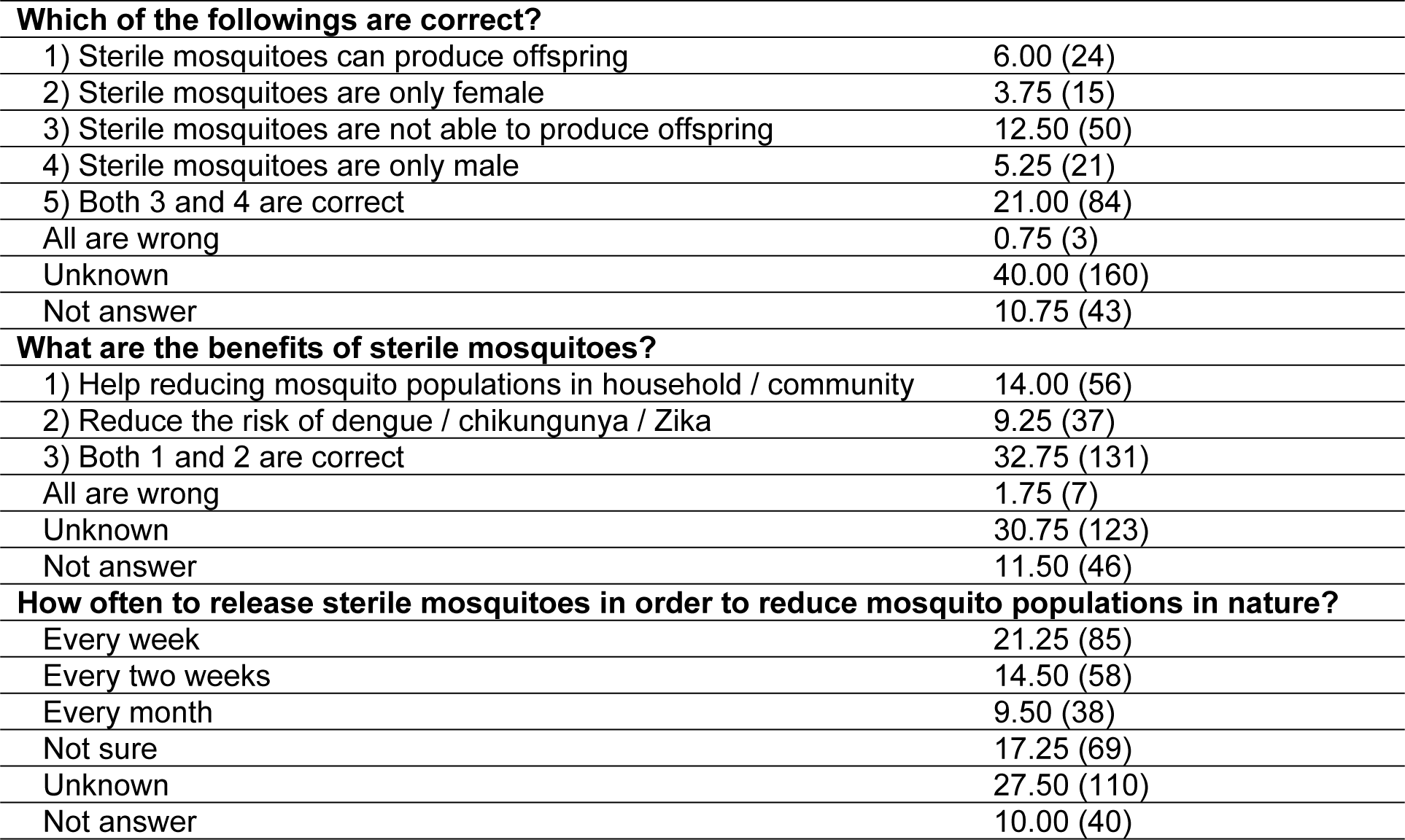
Knowledge on sterile mosquitoes of the surveyed participants living in Bangkok, Thailand.

| Characteristics | % (N = 400) |
| --- | --- |
| **Have you heard about sterilization of mosquitoes to reduce the mosquito vectors of dengue, chikungunya and Zika?** |  |
| Yes | 37.75 (151) |
| No | 45.50 (182) |
| Unknown/Not answer | 16.75 (67) |
| **If yes, where did you receive information about sterilization of mosquitoes in order to reduce the mosquito vectors of dengue, chikungunya, and Zika? (can answer more than one)** |  |
| Local newspapers | 12.57 (24) |
| Village broadcast tower | 13.61 (26) |
| Municipal Officer | 19.90 (38) |
| Billboard | 12.04 (23) |
| Leaflet / Flyer | 16.75 (32) |
| Others | 10.47 (20) |
| Unknown | 14.66 (28) |
| **Do you think you would provide information about mosquito sterilization to your family, acquaintances and neighbors?** |  |
| Yes | 58.50 (234) |
| No | 4.75 (19) |
| Not sure | 14.50 (58) |
| Unknown | 11.75 (47) |
| Not answer | 10.50 (42) |
| **What kinds of mosquitoes are used for sterilization?** |  |
| All kinds of mosquitoes | 24.75 (99) |
| *Aedes* mosquitoes | 32.50 (130) |
| *Culex* mosquitoes | 0.25 (1) |
| *Anopheles* mosquitoes | 1.25 (5) |
| Others | 0 (0) |
| Unknown | 35.00 (140) |
| Not answer | 6.25 (25) |
| **Which of the followings are the methods used to sterilize *Aedes* mosquitoes?** |  |
| Low dose irradiation | 7.50 (30) |
| Injection of bacteria that resist dengue/ chikungunya/ Zika viruses into mosquitoes | 7.00 (28) |
| Both are correct | 29.50 (118) |
| Unknown | 46.50 (186) |
| Not answer | 9.50 (38) |
| **What sex of *Aedes* mosquitoes is used for sterilization?** |  |
| Males | 24.25 (97) |
| Females | 14.75 (59) |
| Both males and females | 19.50 (78) |
| Unknown | 36.50 (146) |
| Not answer | 5.00 (20) |
| **What is the difference between *Aedes* males and females?** |  |
| Males do not feed on blood, only on nectar | 28.25 (113) |
| Females feed on blood, carrying dengue fever | 33.25 (133) |
| Males have thick and long antennae | 8.75 (35) |
| Females do not have thick and long antennae | 0.25 (1) |
| All of the above | 4.50 (18) |
| More than one item | 1.75 (7) |
| Not sure | 5.75 (23) |
| Unknown | 13.75 (55) |
| Not answer | 3.75 (15) |
| **How sterile mosquitoes are different from wild mosquitoes?** |  |
| 1) Sterile mosquitoes are able to resist germs | 11.25 (45) |
| 2) Sterile mosquitoes are unable to mate with wild mosquitoes | 6.25 (25) |
| 3) Sterile mosquitoes can mate with wild mosquitoes but cannot produce offspring | 11.75 (47) |
| 4) Both 1 and 3 are correct | 14.25 (57) |
| 5) All are correct | 10.25 (41) |
| Unknown | 36.25 (145) |
| Not answer | 10.00 (40) |
| **Which of the followings are correct?** |  |
| 1) Sterile mosquitoes can produce offspring | 6.00 (24) |
| 2) Sterile mosquitoes are only female | 3.75 (15) |
| 3) Sterile mosquitoes are not able to produce offspring | 12.50 (50) |
| 4) Sterile mosquitoes are only male | 5.25 (21) |
| 5) Both 3 and 4 are correct | 21.00 (84) |
| All are wrong | 0.75 (3) |
| Unknown | 40.00 (160) |
| Not answer | 10.75 (43) |
| **What are the benefits of sterile mosquitoes?** |  |
| 1) Help reducing mosquito populations in household / community | 14.00 (56) |
| 2) Reduce the risk of dengue / chikungunya / Zika | 9.25 (37) |
| 3) Both 1 and 2 are correct | 32.75 (131) |
| All are wrong | 1.75 (7) |
| Unknown | 30.75 (123) |
| Not answer | 11.50 (46) |
| **How often to release sterile mosquitoes in order to reduce mosquito populations in nature?** |  |
| Every week | 21.25 (85) |
| Every two weeks | 14.50 (58) |
| Every month | 9.50 (38) |
| Not sure | 17.25 (69) |
| Unknown | 27.50 (110) |
| Not answer | 10.00 (40) |

The majority of participants were not able to identify mosquito species to be used for sterilization (35.00%) (Table 6) but some were able to identify *Aedes* mosquitoes as the species to be used for sterilization (32.50%). However, some of them stated that all kinds of mosquitoes could be sterilized (24.75%) (Table 6). The majority of people did not know how to make mosquitoes sterile (46.50%) but some believed both low dose radiation and injection of bacteria resistant to dengue, chikungunya, and Zika virus into mosquitoes (29.50%) could be used to produce sterile mosquitoes. The majority of people did not know about what sex of mosquitoes to be selected for sterilization (36.50%). Some believed only males (24.25%), some believed only females (14.15%), and some believed both sexes could be sterilized (19.50%).

The majority of participants were able to differentiate the behavior of *Aedes* males and females. 61.50% was able to identify that females fed on blood meals, and males only fed on nectar (Table 6). When asked about sterile mosquitoes, the majority of people was able to differentiate sterile from wild mosquitoes (53.75%), i.e., sterile mosquitoes were able to resist germs, and they can mate with wild mosquitoes but cannot produce their offspring; but some people were still uncertain or unknown (19.50%). However, there was a big gap of information between those who know and those who did not know about sterile mosquitoes in this study. The majority of people still did not know any information about sterile mosquitoes (40.00%), whereas some of them had very well and correct information about sterile mosquitoes (38.75%). This fact made many people still did not realize any benefits of sterile mosquitoes (30.75%). However, some of them already knew that sterile mosquitoes could help reducing diseases and mosquito populations (56.00%).

### Attitude towards the application of sterile mosquitoes

The majority of people had positive attitude towards sterile mosquitoes and many people agreed or strongly agreed that application of sterile mosquitoes could be effective, practical and safe for human, animals, and environment (47.00%). However, some of them were still uncertain on this aspect (29.00%) (Table 7). Many people believed that sterile mosquitoes could reduce mosquito vectors of dengue, chikungunya, and Zika (35.25%) but slightly lower percentage of people were still uncertain (33.00%). The majority of people believed that the application of sterile mosquitoes was more practical than the use of chemicals to reduce mosquito vectors (32.50%). However, many of them were still uncertain (27.00%) and some of them believed that it was less effective (2.00%) or even useless (2.25%).

**Table 7.** Attitude towards the application of sterile mosquitoes of surveyed participants living in Bangkok, Thailand.

| Characteristics | % (N = 400) |
| --- | --- |
| **Do you think sterile mosquitoes are effective, practical and safe for human, animals and environment?** |  |
| Strongly agree | 17.25 (69) |
| Agree | 29.75 (119) |
| Not sure | 29.00 (116) |
| Disagree | 1.00 (4) |
| Strongly disagree | 0.75 (3) |
| Do not know / Do not answer | 22.25 (89) |
| **Do you think sterile mosquitoes can be used to reduce mosquito vectors of dengue, chikungunya and Zika?** |  |
| Yes | 35.25 (141) |
| No | 7.25 (29) |
| Not sure | 33.00 (132) |
| Do not know/ Do not answer | 24.50 (98) |
| **Do you think an application of sterile mosquitoes is more useful than chemicals in order to reduce the mosquito vectors of dengue, chikungunya and Zika?** |  |
| More useful | 32.50 (130) |
| Equally | 8.75 (35) |
| Less useful | 2.00 (8) |
| Useless | 2.25 (9) |
| Not sure | 27.00 (108) |
| Do not know/ Do not answer | 27.50 (110) |
| **If an application of sterile mosquitoes can reduce the mosquito vectors of dengue, chikungunya and Zika, would you like to have the sterile mosquitoes introduced into your household or community?** |  |
| Desperately need | 19.75 (79) |
| Need | 25.50 (102) |
| Not sure | 17.75 (71) |
| Not require | 3.50 (14) |
| Absolutely not need | 0.50 (2) |
| Do not know/ Do not answer | 33.00 (132) |
| **Could you please give rating on the application of sterile mosquitoes in order to reduce the mosquito vectors of dengue, chikungunya and Zika?** |  |
| Absolutely not good | 1.25 (5) |
| Not good | 7.00 (28) |
| Moderate | 29.75 (119) |
| Good | 29.00 (116) |
| Very good | 23.75 (95) |
| Do not know/ Do not answer | 9.25 (37) |
| **Are you interested in implementing new technologies or methods to reduce the mosquito** |  |

| **vectors of dengue, chikungunya and Zika in your household or community?** |  |
| --- | --- |
| Extremely interested | 24.50 (98) |
| Interested | 38.25 (153) |
| Not sure | 21.00 (84) |
| Not interested | 1.25 (5) |
| Strongly not interested | 0.50 (2) |
| Do not know/ Do not answer | 14.50 (58) |
| **What factors influence your decision to adopt new technologies or methods to reduce dengue, chikungunya, and Zika in your homes or communities? (can answer more than one)** |  |
| Price and cost effectiveness | 14.00 (56) |
| Effectiveness | 18.50 (74) |
| Human safety | 24.25 (97) |
| Animal safety | 2.75 (11) |
| Environmental safety | 13.00 (52) |
| Others | 0.25 (1) |
| Do not know/ Do not answer | 27.25 (109) |

When asked whether participants would like to have sterile mosquitoes applied into their homes or communities, the majority of them desperately wanted or wanted (45.25%), while some people did not reply (33.00%), some were still uncertain (17.75%), and some did not required or absolutely not needed (4.00%). When asked people to rate overall application of sterile mosquitoes, the majority of people rated as good to very good (52.75%), followed by moderate (29.75%), did not answer (9.25%), and bad to very bad (8.25%). When asked people about their interest in implementing sterile mosquitoes in their homes or communities, the majority of them were interested or extremely interested (62.75%) followed by uncertain (21.00%), did not answer (14.50%), and only a few of them was not interested or strongly not interested (1.75%). When asked about factors that might influence their decision of using sterile mosquitoes in their homes or communities, the majority of them did not answer (27.25%), followed by human safety (24.25%), animal safety (18.50%), price and cost effectiveness (14.00%), and environmental safety (13.00%).

### Health seeking behavior

The majority of surveyed participants primarily had home treatment, i.e., use home - based medications or creams, when they or their family member get bitten by mosquitoes (Table 8). In the case of the participants, after home treatment (29.61%), they preferred to buy medicines or products for their own use (26.34%), followed by going to see a medical doctor (18.75%), or see a specialist (10.12%). However, some of them did nothing (10.12%). In case of their family members, they firstly used home treatment (39.48%), followed by going to see a medical doctor (22.08%), buying products for their own use (10.39%), but some of them did nothing (10.39%).

**Table 8.**
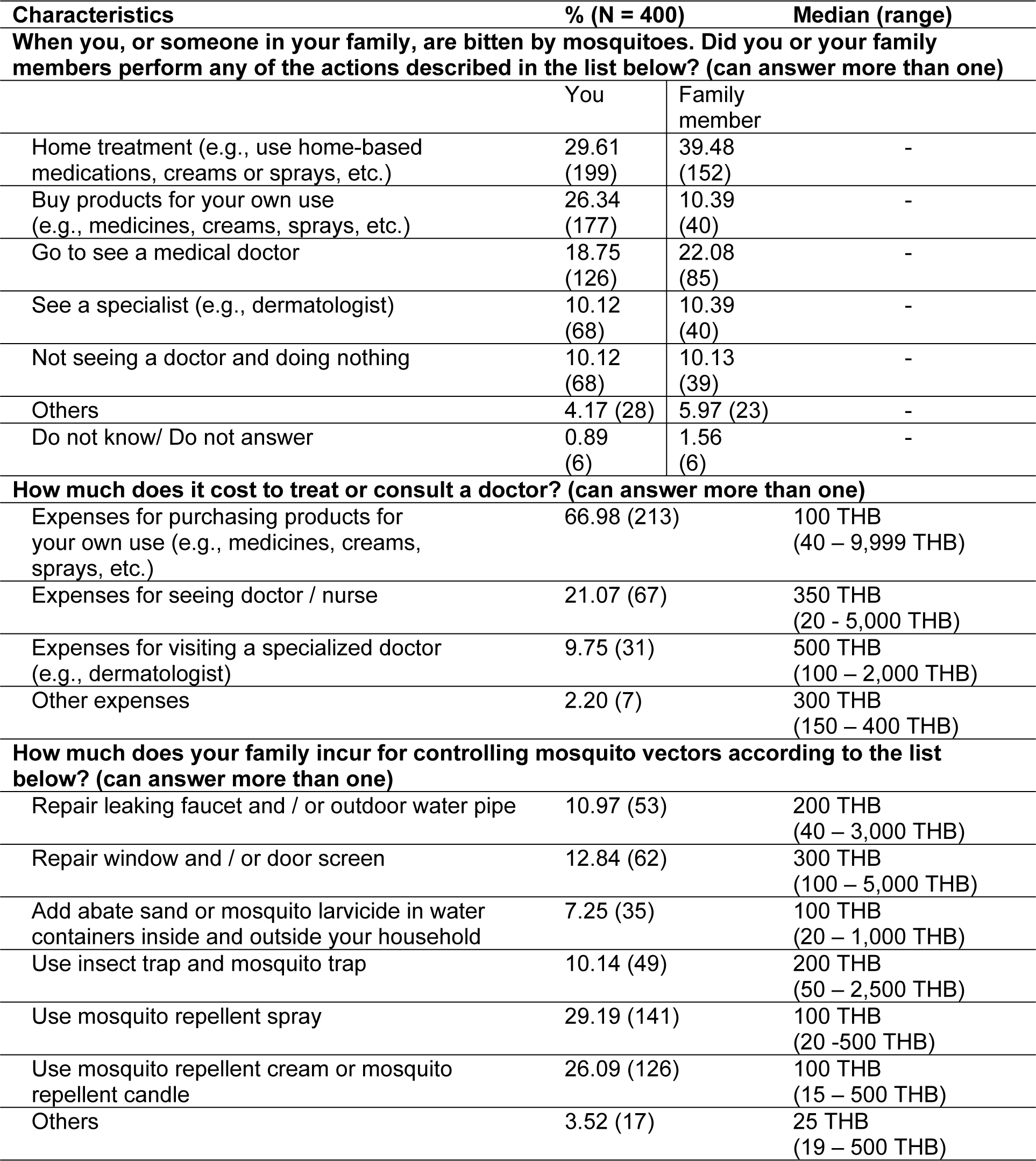
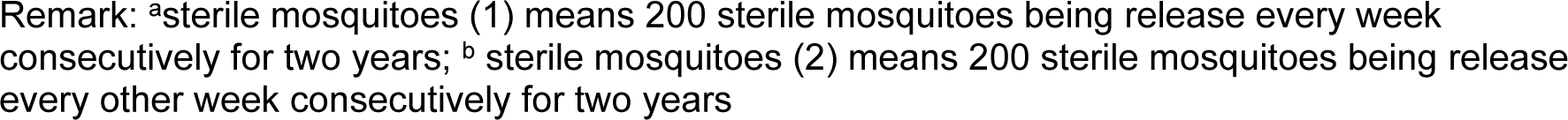
Health seeking behavior of surveyed participants living in Bangkok, Thailand.

When asked about the expense for treatment, the major expense was for purchasing products for their own use, i.e., medicine, creams, sprays of ∼100 THB (66.98%), followed by expenses for seeing doctors of ∼350 THB (21.07%), expenses for visiting specialized doctors of ∼500 THB (9.75%), and other expenses of ∼300 THB (2.20%) (Table 8). Sometimes, expenses could be up to more than 5,000 THB per treatment which caused an economic burden to people since average daily income for Thai people was about 300 THB and the expense for treatment was almost more than their daily incomes without considering the loss of income due to sickness that could be increased when the number of days absent from work increased, especially when they were laborer that rely on the daily income.

When asked about other expenses for vector control, the majority of people spent ∼100 THB for mosquito repellent spray (29.29%) (Table 8), followed by 100 THB for mosquito repellent cream or mosquito repellent candles (26.09%), 300 THB for repairing window or door screens (12.84%), 200 THB for repairing leaking faucets and / or outdoor water pipes (10.97%), 200 THB for insect traps and mosquito traps (10.14%). Sometimes, expense was up to 2,500-5,000 THB if it was for mosquito traps, door screens, leaking faucets or water pipes.

### Willingness to pay for the application of sterile mosquitoes in order to control dengue, chikungunya, and Zika

When asked participants to select one of sterile mosquitoes when it was at their own expenses, the majority of people did not select any options (52.00%), some of them preferred sterile mosquitoes (2) with two-week release frequency (17.50%), followed by sterile mosquitoes (1) with one-week release frequency (10.75%) (Table 9). For those participants who selected sterile mosquitoes (1) with one-week release frequency, the majority of them were not willing to purchase any sterile mosquitoes when they were available on the market (32.75%), only a few of them were willing to pay from 2.50 – 15.00 THB (6.50 – 9.25%), and the maximum that they could pay was 1 THB per one sterile mosquito (9.50%). When asked about the reasons why they did not want to purchase sterile mosquitoes, the majority of them expected to get it for free from the government (14.75%), followed by the reasons that they could not afford to buy (10.50%), or they needed other measures to prevent and control dengue, chikungunya and Zika (6.50%).

**Table 9.** Willingness to pay for the application of sterile mosquitoes to reduce vectors of dengue, chikungunya, and Zika of the surveyed participants living in Bangkok, Thailand.

| Characteristics | % (N = 400) |  |
| --- | --- | --- |
| **Which of the following methods will you choose if the cost is at your own expense?** |  |  |
| Sterile mosquitoes (1)<sup>a</sup> | 10.75 (43) |  |
| Sterile mosquitoes (2)<sup>b</sup> | 17.50 (70) |  |
| Do not select both | 52.00 (208) |  |
| Do not know/ Do not answer | 19.75 (79) |  |
| **If the sterile mosquitoes are sold in the market, are you willing to pay for them?** | **Sterile mosquitoes (1)** | **Sterile mosquitoes (2)** |
| Yes | 12.00 (48) | 12.00 (48) |
| No | 32.75 (131) | 39.50 (158) |
| Do not know/ Do not answer | 55.25 (221) | 48.50 (194) |
| **Are you willing to pay 5 THB for one sterile mosquito?** |  |  |
| Yes | 9.25 (37) | 9.50 (38) |
| No | 29.00 (116) | 29.75 (119) |
| Do not know/ Do not answer | 61.75 (247) | 60.75 (243) |
| **Are you willing to pay 10 THB one sterile mosquito?** |  |  |
| Yes | 7.50 (30) | 7.25 (29) |
| No | 29.75 (119) | 27.25 (109) |
| Do not know/ Do not answer | 62.75 (251) | 65.50 (262) |
| **Are you willing to pay 15 THB one sterile mosquito?** |  |  |
| Yes | 6.50 (26) | 6.50 (26) |
| No | 28.75 (115) | 28.50 (114) |
| Do not know/ Do not answer | 64.75 (259) | 65.00 (260) |
| **Are you willing to pay 2.5 THB one sterile mosquito?** |  |  |
| Yes | 7.00 (28) | 6.50 (26) |
| No | 29.50 (118) | 29.50 (118) |
| Do not know/ Do not answer | 63.50 (254) | 64.00 (256) |
| **What is the maximum amount you are willing to pay for each sterile mosquito?** | 9.50 (38)<br>median (range)<br>1 THB<br>(1-250 THB) | 9.00 (36)<br>median (range)<br>2 THB<br>(1 – 200 THB) |
| Do not know/ Do not answer | 90.50 (362) | 91.00 (364) |
| **Please indicate the reason why you refuse to pay for sterile mosquitoes.** |  |  |
| Want to get sterile mosquitoes free from the Government | 14.75 (59) | 37.75 (151) |
| Want to know more information or scientific evidence about sterile mosquitoes | 3.50 (14) | 13.00 (52) |
| Cannot afford to buy sterile mosquitoes | 10.50 (42) | 17.75 (71) |
| Need other measures to prevent and control dengue, chikungunya and Zika | 6.50 (26) | 9.75 (39) |
| Others | 1.25 (5) | 2.25 (9) |
| Do not know/ Do not answer | 63.25 (253) | 19.50 (78) |

For those who selected sterile mosquitoes (2) with two-week release frequency, the majority of them also did not willing to purchase any sterile mosquitoes when they were available on the market (39.50%), only a few of them were willing to pay from 2.50 – 15.00 THB (6.50 – 9.50%), and the maximum that they could pay was 2 THB per one sterile mosquito (9.00%), which was almost double from those of sterile mosquitoes (1) with one-week release frequency (Table 9). When asked about the reasons why they did not want to purchase sterile mosquitoes, the majority of them also wanted to get it for free from the government (36.75%), which was much higher percentage than those who selected sterile mosquitoes (1) with one-week release frequency, followed by the reasons that they could not afford to buy (17.75%), they wanted to know more information or scientific evidence (13.00%), or they needed other measures to prevent and control dengue, chikungunya and Zika (9.75%). For comparison, for those who selected sterile mosquitoes (2) with two-week release frequency, they wanted to know more information in much higher percentage than those who selected sterile mosquitoes (1) with one-week release frequency (13.00% vs 3.50%).

### Impact on environment, economic, social, and quality of life from the release of sterile mosquitoes

The majority of participants were positive when asked about environmental, economic, social impacts including the impact on their quality of life from the release of sterile mosquitoes. In term of the environmental impact, the majority of people agreed or strongly agreed that release of sterile mosquitoes could eliminate the breeding sites of mosquito larvae (81.50%), they believed it could lead to a better waste management (82.50%), they could have cleaner home or community or pleasant environment (82%), they could make use of mosquito catching equipment/tools in their homes or communities (72.25%), the number of mosquitoes in the household or community decreased (68%), chemical usage or residual pollution could be reduced from controlling mosquitoes (70.50%), and their homes or communities could be free from dengue, chikungunya and Zika (71.00%) (Table 10). However, higher percentage of uncertainty was increased from 16.25-17.25%, and the percentage of disagree to strongly disagree was also increased from 4.25-5.25% when asked people about reduction of mosquitoes, reduction of chemical use, and home or community free from dengue, chikungunya and Zika when compared to other aspects when the percentage of disagree or strongly disagree was low at only 0.75-2.25%.

**Table 10.** Impacts on the environment resulting from the release of sterile mosquitoes of the surveyed participants living in Bangkok, Thailand.

| Characteristics | % (N = 400) |
| --- | --- |
| **Environmental impacts** |  |
| **Breeding sites of mosquito larvae are eliminated in household / community** |  |
| Totally agree | 48.00 (192) |
| Agree | 33.50 (134) |
| Uncertain | 7.00 (28) |
| Disagree | 2.25 (9) |
| Strongly disagree | 0 (0) |
| Do not know / Do not answer | 9.25 (37) |
| **Better waste management in household / community. Being waste-free or good hygienic communities** |  |
| Totally agree | 46.50 (186) |
| Agree | 36.00 (144) |
| Uncertain | 7.50 (30) |
| Disagree | 1.75 (7) |
| Strongly disagree | 0 (0) |
| Do not know / Do not answer | 8.25 (33) |
| **Cleaner household / community and pleasant environment** |  |
| Totally agree | 47.75 (191) |
| Agree | 34.25 (137) |
| Uncertain | 8.25 (33) |
| Disagree | 1.00 (4) |
| Strongly disagree | 0.50 (2) |
| Do not know / Do not answer | 8.25 (33) |
| **Can make use of mosquito catching equipment / tools in household / community** |  |
| Totally agree | 33.50 (134) |
| Agree | 38.75 (155) |
| Uncertain | 17.75 (71) |
| Disagree | 0.25 (1) |
| Strongly disagree | 0.50 (2) |
| Do not know / Do not answer | 9.25 (37) |
| **Number of mosquitoes in household / community decreases** |  |
| Totally agree | 35.50 (142) |
| Agree | 32.50 (130) |
| Uncertain | 16.75 (67) |
| Disagree | 3.25 (13) |
| Strongly disagree | 1.50 (6) |
| Do not know / Do not answer | 10.50 (42) |
| **Reduce the use of chemicals to control mosquitoes. Reduce residual pollution resulting from the use of chemicals in the environment** |  |
| Totally agree | 39.25 (157) |
| Agree | 31.25 (125) |
| Uncertain | 16.25 (65) |
| Disagree | 3.50 (14) |
| Strongly disagree | 1.75 (7) |
| Do not know / Do not answer | 8.00 (32) |
| **Household / community is free from dengue, chikungunya and Zika** |  |
| Totally agree | 38.25 (153) |
| Agree | 32.75 (151) |
| Uncertain | 17.25 (69) |
| Disagree | 2.75 (11) |
| Strongly disagree | 1.50 (6) |
| Do not know / Do not answer | 7.50 (30) |

In terms of an economic impact, the majority of people agreed or strongly agreed that release of sterile mosquitoes could reduce household or community expenses in purchasing mosquito repellent (68.75%), reducing the cost of medical treatment (71.25%), less or no loss of income due to the absence from work (70.50%), having more savings without medical expenses due to illness (69.75%), having more income from being volunteered to collect mosquito samples in the area (64.25%). In addition, they believed that the community shops could have more income as researcher come to do research in the area (55.75%) (Table 11). However, more percentage of uncertainty was observed when asked about more income from research activities or researcher (25.50-32.35%).

**Table 11.**
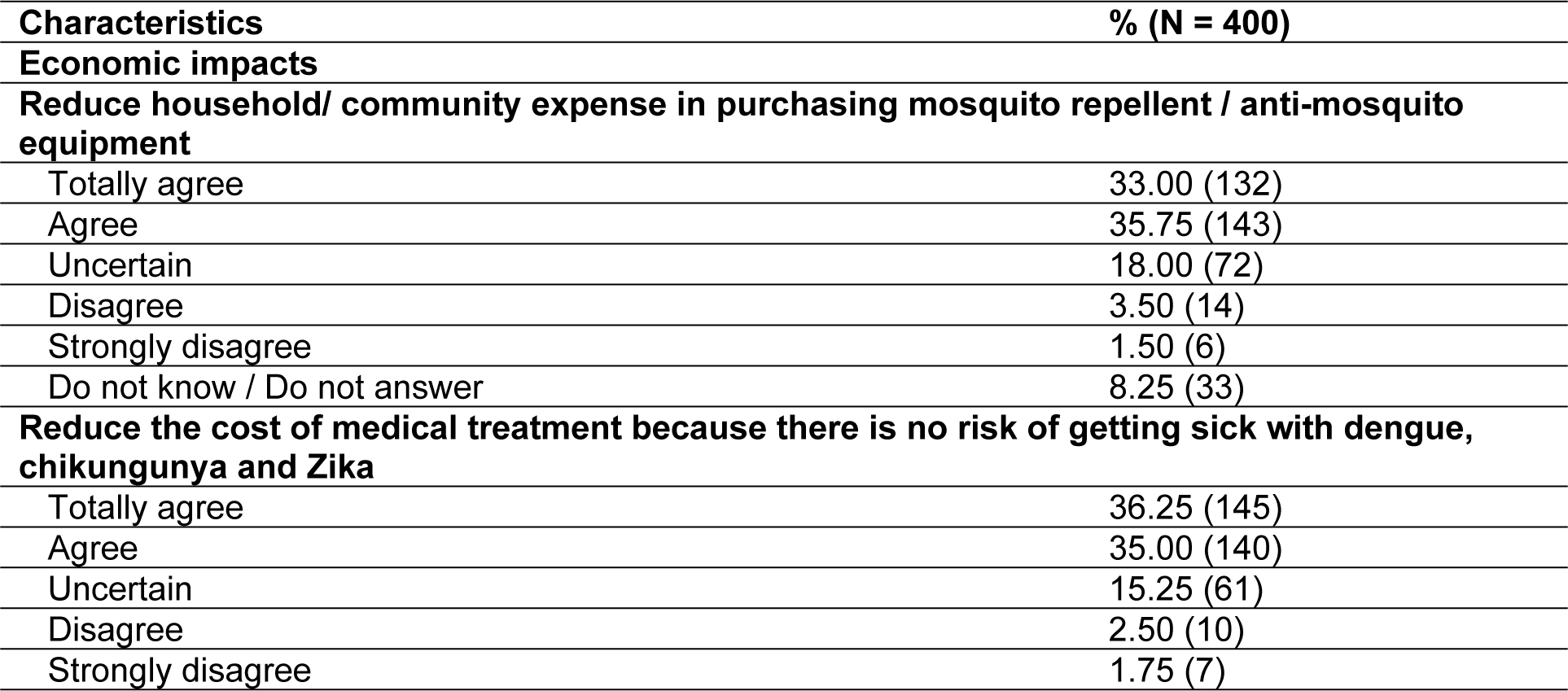
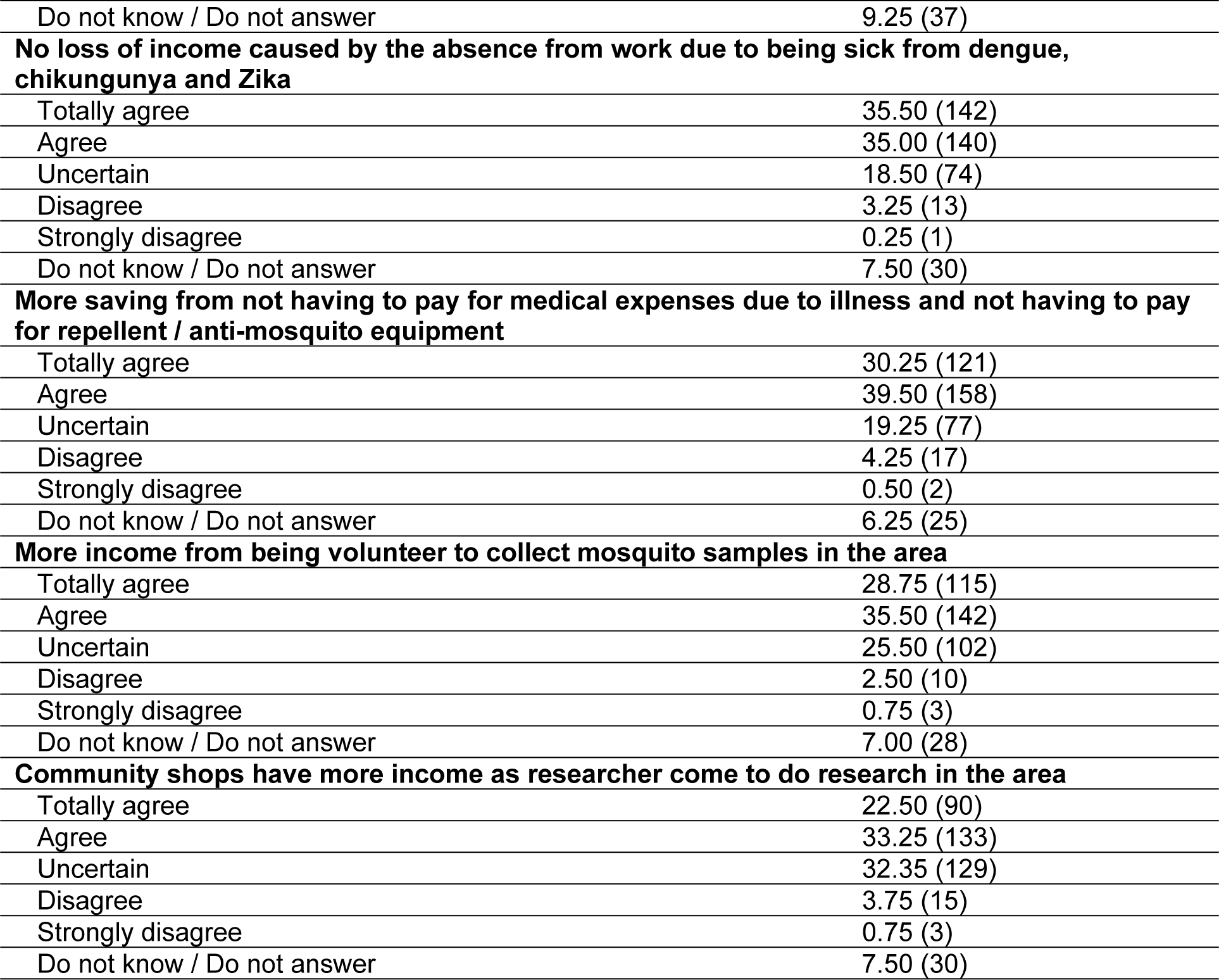
Economic impacts resulting from the release of sterile mosquitoes of the surveyed participants living in Bangkok, Thailand.

| **Characteristics** | **% (N = 400)** |
| --- | --- |
| **Economic impacts** |  |
| **Reduce household/ community expense in purchasing mosquito repellent / anti-mosquito equipment** |  |
| Totally agree | 33.00 (132) |
| Agree | 35.75 (143) |
| Uncertain | 18.00 (72) |
| Disagree | 3.50 (14) |
| Strongly disagree | 1.50 (6) |
| Do not know / Do not answer | 8.25 (33) |
| **Reduce the cost of medical treatment because there is no risk of getting sick with dengue, chikungunya and Zika** |  |
| Totally agree | 36.25 (145) |
| Agree | 35.00 (140) |
| Uncertain | 15.25 (61) |
| Disagree | 2.50 (10) |
| Strongly disagree | 1.75 (7) |
| Do not know / Do not answer | 9.25 (37) |
| **No loss of income caused by the absence from work due to being sick from dengue, chikungunya and Zika** |  |
| Totally agree | 35.50 (142) |
| Agree | 35.00 (140) |
| Uncertain | 18.50 (74) |
| Disagree | 3.25 (13) |
| Strongly disagree | 0.25 (1) |
| Do not know / Do not answer | 7.50 (30) |
| **More saving from not having to pay for medical expenses due to illness and not having to pay for repellent / anti-mosquito equipment** |  |
| Totally agree | 30.25 (121) |
| Agree | 39.50 (158) |
| Uncertain | 19.25 (77) |
| Disagree | 4.25 (17) |
| Strongly disagree | 0.50 (2) |
| Do not know / Do not answer | 6.25 (25) |
| **More income from being volunteer to collect mosquito samples in the area** |  |
| Totally agree | 28.75 (115) |
| Agree | 35.50 (142) |
| Uncertain | 25.50 (102) |
| Disagree | 2.50 (10) |
| Strongly disagree | 0.75 (3) |
| Do not know / Do not answer | 7.00 (28) |
| **Community shops have more income as researcher come to do research in the area** |  |
| Totally agree | 22.50 (90) |
| Agree | 33.25 (133) |
| Uncertain | 32.35 (129) |
| Disagree | 3.75 (15) |
| Strongly disagree | 0.75 (3) |
| Do not know / Do not answer | 7.50 (30) |

In terms of a social impact, the majority of people agreed or strongly agreed that release of sterile mosquitoes could provide them more opportunity to discuss or to take responsibility on elimination of mosquito breeding sites with their family members (70.50%), they could get along well with neighbors (72.50%), they could have more opportunity to communicate with neighbors about mosquito control activities (67.00%), they could receive health support from community volunteers or health officers (73.50%), they were willing to assist their neighbors or community in eliminating mosquito breeding sites (74.00%), they could have a good relationship with neighbors or people in the community (58.25%), they were satisfied with the living conditions (59.75%), and they were better aware of various information from media, including community broadcast channel (62.00%) (Table 12). However, high percentage of uncertainty was observed when asked about good relationship with neighbors, satisfaction of living condition, and awareness of information from the release of sterile mosquitoes (22.00-27.75%).

**Table 12.** Social impacts resulting from the release of sterile mosquitoes of the surveyed participants living in Bangkok, Thailand.

| Characteristics | % (N = 400) |
| --- | --- |
| **Social Impacts** |  |
| **You discuss, consult and take responsibility about the elimination of mosquito breeding sites and mosquito eradication with your family members.** |  |
| Totally agree | 25.75 (103) |
| Agree | 44.75 (179) |
| Uncertain | 17.75 (71) |
| Disagree | 3.00 (12) |
| Strongly disagree | 0.25 (1) |
| Do not know / Do not answer | 8.50 (34) |
| **You get along well with your neighbors/people in the community because you participate in community activities.** |  |
| Totally agree | 27.50 (110) |
| Agree | 45.00 (180) |
| Uncertain | 17.00 (68) |
| Disagree | 2.50 (10) |
| Strongly disagree | 0.50 (2) |
| Do not know / Do not answer | 7.50 (30) |
| **You have more opportunity to communicate with neighbors / people in the community about mosquito control activities.** |  |
| Totally agree | 23.75 (95) |
| Agree | 43.25 (173) |
| Uncertain | 22.00 (88) |
| Disagree | 3.25 (13) |
| Strongly disagree | 0.25 (1) |
| Do not know / Do not answer | 7.50 (30) |
| **You receive health support / care / promotion from community volunteers / health officials.** |  |
| Totally agree | 28.50 (114) |
| Agree | 45.00 (180) |
| Uncertain | 16.50 (66) |
| Disagree | 2.50 (10) |
| Strongly disagree | 0.50 (2) |
| Do not know / Do not answer | 7.00 (28) |
| **You are willing to assist your neighbors / community in eliminating mosquito breeding sites and controlling mosquitoes.** |  |
| Totally agree | 31.00 (124) |
| Agree | 43.00 (172) |
| Uncertain | 16.25 (65) |
| Disagree | 3.25 (13) |
| Strongly disagree | 0.25 (1) |
| Do not know / Do not answer | 6.25 (25) |
| **After releasing sterile mosquitoes, you and people within the community have good relationship.** |  |
| Totally agree | 24.25 (97) |
| Agree | 34.00 (136) |
| Uncertain | 27.25 (109) |
| Disagree | 4.00 (16) |
| Strongly disagree | 1.75 (7) |
| Do not know / Do not answer | 8.75 (35) |
| **You are satisfied with the living conditions after conducting a research project in the area** |  |
| Totally agree | 22.75 (91) |
| Agree | 37.00 (148) |
| Uncertain | 27.75 (111) |
| Disagree | 3.00 (12) |
| Strongly disagree | 0.50 (2) |
| Do not know / Do not answer | 9.00 (36) |
| **You are better aware of various information from printed media, radio, television and wired community news** |  |
| Totally agree | 21.75 (87) |
| Agree | 40.25 (161) |
| Uncertain | 26.00 (104) |
| Disagree | 2.75 (11) |
| Strongly disagree | 0.75 (3) |
| Do not know / Do not answer | 8.50 (34) |

In terms of an impact on the quality of life, the majority of people agreed or strongly agreed that release of sterile mosquitoes could make them safe and satisfied with life (70.00%), they could have good mental health with no stress related to illness (70.50%), they felt to have more self-valued due to their contribution to the reduction of mosquito breeding sites in the community (69.75%), they could take a break from the stress due to unpleasant community environment (66.50%), and they or their communities could gain benefits from the release of sterile mosquitoes (61.25%) (Table 13). However, people were more uncertained when asked about an opportunity to take a break from stress environment (20.75%) or the benefits that they could get from the release of sterile mosquitoes (26.75%).

**Table 13.** Impacts on the quality of life resulting from the release of sterile mosquitoes of the surveyed participants living in Bangkok, Thailand.

| Characteristics | % (N = 400) |
| --- | --- |
| **Impacts on the quality of life** |  |
| **You feel safe and satisfied with your life because you are not afraid of getting sick with mosquito-borne diseases?** |  |
| Totally agree | 28.50 (114) |
| Agree | 41.50 (166) |
| Uncertain | 18.50 (74) |
| Disagree | 1.75 (7) |
| Strongly disagree | 0.25 (1) |
| Do not know / Do not answer | 9.50 (38) |
| **You are in good mental health with no stress due to good health and no illness** |  |
| Totally agree | 29.25 (117) |
| Agree | 41.25 (165) |
| Uncertain | 17.25 (69) |
| Disagree | 2.25 (9) |
| Strongly disagree | 0.25 (1) |
| Do not know / Do not answer | 9.75 (39) |
| **You feel to have more self-valued because of your contribution to reduce mosquito breeding sites / eradicating mosquitoes in your household / community?** |  |
| Totally agree | 30.00 (120) |
| Agree | 39.75 (159) |
| Uncertain | 17.50 (70) |
| Disagree | 1.50 (6) |
| Strongly disagree | 0.50 (2) |
| Do not know / Do not answer | 10.75 (43) |
| **You can take a break from stress due to unpleasant community environment** |  |
| Totally agree | 27.00 (108) |
| Agree | 39.50 (158) |
| Uncertain | 20.75 (83) |
| Disagree | 2.00 (8) |
| Strongly disagree | 0.25 (1) |
| Do not know / Do not answer | 10.50 (42) |
| **You and your community can gain benefits from the release of sterile mosquitoes** |  |
| Totally agree | 25.75 (103) |
| Agree | 35.50 (142) |
| Uncertain | 26.50 (106) |
| Disagree | 1.75 (7) |
| Strongly disagree | 0.25 (1) |
| Do not know / Do not answer | 10.25 (41) |

### Odds ratios of the knowledge on dengue with socio-demographic characteristics

In this study, we could identify some significant correlations between the knowledge on dengue and socio-demographic characteristics as shown in Table 14. Firstly, we found that the participants who were tenants had significantly 2.84 times higher knowledge on dengue transmission that it was through mosquito bites than those who were owners of the houses (OR = 2.84, 95% CI = 1.04 – 7.77, *p* = 0.041). When focused on the biting time of *Aedes* mosquitoes, we found that participants aged more than 54 years had significantly 41% lower knowledge than those aged below 35 years (OR = 0.59, 95% CI = 0.35 – 0.99, *p* = 0.047). For participants with monthly incomes between 5,001-10,000 THB (US$ 144-289), they had significantly 4.57 times higher knowledge on the biting time of *Aedes* mosquitoes when compared to those with the incomes lower than 5,000 THB (OR = 4.57, 95% CI = 1.33 – 15.67, *p* = 0.016). When focused on previous dengue virus infections, participants who were spouses (OR = 0.27, 95% CI = 0.09 – 0.82, *p* = 0.021) and relatives (OR = 0.16, 95% CI = 0.04 – 1.24, *p* = 0.019) had significantly 73% and 84% lower experience of getting historical dengue virus infections respectively than those who were heads of the family. In addition, participants who were tenants had significantly 1.83 times higher experience of getting dengue infections when compared to those who were owners of the house (OR = 1.83, 95%CI = 1.07 – 3.15, p = 0.028).

**Table 14.** Odds ratios of the knowledge on dengue with socio-demographic characteristics.

| Variable | Odd Ratios | 95%CI | P-value |
| --- | --- | --- | --- |
| **I. Dengue is transmitted through mosquito bites** |  |  |  |
| **House ownership** |  |  |  |
| Owner | Ref |  |  |
| Tenant | 2.84 | 1.04 – 7.77 | 0.042* |
| **II. Day biting time of *Aedes* mosquitoes** |  |  |  |
| **Age** |  |  |  |
| Age < 35 | Ref |  |  |
| Age 35-54 | 0.61 | 0.31 – 1.21 | 0.158 |
| Age >54 | 0.59 | 0.35 – 0.99 | 0.047* |
| **Monthly income (THB) (US\$ 1 = 34 THB)** | | | |
| < 5,000 | Ref |  |  |
| 5,001-10,000 | 4.57 | 1.33 – 15.67 | 0.016* |
| 10,001-15,000 | 1.70 | 0.76 – 3.82 | 0.198 |
| 15,001-20,000 | 1.24 | 0.53 – 2.89 | 0.620 |
| 20,001-30,000 | 1.81 | 0.73 – 4.48 | 0.202 |
| > 30,000 | 0.85 | 0.30 – 2.42 | 0.758 |
| **III. Experience of the historical dengue virus infections** |  |  |  |
| **Family status** |  |  |  |
| Head of family | Ref |  |  |
| Spouse | 0.27 | 0.09 – 0.82 | 0.021* |
| Son/daughter | 0.60 | 0.20 – 1.80 | 0.360 |
| Relatives | 0.16 | 0.03 – 0.74 | 0.019* |
| Others | 0.21 | 0.04 – 1.24 | 0.085 |
| **House ownership** |  |  |  |
| Owner | Ref |  |  |
| Tenant | 1.83 | 1.07 – 3.15 | 0.028* |

\* Significant different at $p < 0.05$

### Odds ratios of the knowledge on chikungunya with socio-demographic characteristics

For chikungunya, we found that family status, occupation, and number of family members had significant correlations with knowledge on chikungunya transmission. Participants who were spouses had significantly 4.37 times higher knowledge on chikungunya transmission when compared to those who were heads of the family (OR = 4.37, 95% CI = 1.21 – 15.78, *p* = 0.024) (Table 15). Merchants had significantly 5.65 times higher knowledge than those of laborers (OR = 5.65, 95% CI = 1.39 – 23.05, *p* = 0.016); and those who lived with more than 5 people in the family significantly had 58% lower knowledge than those living with 1-2 family members (OR = 0.42, 95% CI = 0.18 – 0.97, *p* = 0.042). In term of the biting time of *Aedes* mosquitoes, the family status, education, and occupation had significant correlations with knowledge on this variable. Participants who were sons or daughters had significantly 3.97 times higher knowledge than those who were heads of the family (OR = 3.97, 95% CI = 1.12 – 14.14, *p* = 0.032). Participants with the highest education at secondary school had significantly 1.11 times higher knowledge than those with no education (OR = 1.11, 95% CI = 1.11 – 24.48, *p* = 0.036). Lastly, merchants or other occupations had significantly 3.42 (OR = 3.42, 95% CI = 1.25 – 9.31, *p* = 0.016) and 4.02 (OR = 4.02, 95% CI = 1.28 – 12.65, *p* = 0.017) times higher knowledge than those who worked as laborers respectively.

**Table 15.** Odds ratios of the knowledge on chikungunya and sterile mosquitoes with socio-demographic characteristics.

| Variable | Odd Ratios | 95%CI | P-value |
| --- | --- | --- | --- |
| **I. Chikungunya is transmitted through mosquito bites** |  |  |  |
| **Family status** |  |  |  |
| Head of family | Ref |  |  |
| Spouse | 2.72 | 0.83 – 8.92 | 0.098 |
| Son/daughter | 4.37 | 1.21 – 15.78 | 0.024* |
| Relative | 3.43 | 0.73 – 16.09 | 0.118 |
| Others | 2.57 | 0.37 – 17.83 | 0.339 |
| **Occupation** |  |  |  |
| Laborer | Ref |  |  |
| Farmer | - | - | - |
| Merchant | 5.65 | 1.39 – 23.05 | 0.016* |
| Government/<br>State Enterprise officer | 1.39 | 0.41 – 4.72 | 0.603 |
| Company employer | 0.80 | 0.26 – 2.46 | 0.699 |
| Housekeeper | 2.77 | 0.27 – 28.39 | 0.391 |
| Others | 1.56 | 0.45 – 5.43 | 0.486 |
| **Number of family members** |  |  |  |
| 1 - 2 | Ref |  |  |
| 3 – 4 | 0.62 | 0.23 – 1.69 | 0.349 |
| > 5 | 0.42 | 0.18 – 0.97 | 0.042* |
| **II. Biting time of *Aedes* mosquitoes** |  |  |  |
| **Family status** |  |  |  |
| Head of family | Ref |  |  |
| Spouse | 2.26 | 0.70 -7.28 | 0.171 |
| Son/daughter | 3.97 | 1.12 – 14.14 | 0.032* |
| Relative | 1.46 | 0.39 – 5.51 | 0.578 |
| Others | 0.73 | 0.15 – 3.47 | 0.692 |
| **Education** |  |  |  |
| None | Ref |  |  |
| Primary school | 5.83 | 0.70 – 48.87 | 0.104 |
| Secondary school | 1.11 | 1.11 – 24.48 | 0.036* |
| High school | 0.77 | 0.77 – 17.64 | 0.102 |
| Bachelor degree | 0.48 | 0.48 – 11.30 | 0.292 |
| Higher than Bachelor degree | 0.44 | 0.44 – 9.30 | 0.370 |
| **Occupation** |  |  |  |
| Laborer | Ref |  |  |
| Farmer | - | - | - |
| Merchant | 3.42 | 1.25 – 9.31 | 0.016* |
| Government/<br>State Enterprise officer | 1.87 | 0.64 – 5.51 | 0.256 |
| Company employer | 1.16 | 0.46 – 2.95 | 0.758 |
| Housekeeper | 2.17 | 0.33 – 14.06 | 0.418 |
| Others | 4.02 | 1.28 – 12.65 | 0.017* |

\* Significant difference at $p < 0.05$

### Odds ratios of the knowledge on Zika with socio-demographic characteristics

For Zika, we found a significant correlation between age and occupation with the knowledge on Zika transmission. Participants aged 35-54 years, had significantly 70% lower knowledge than those aged lower than 35 years (OR = 0.30, 95%CI = 0.12 – 0.77, *p* = 0.012); and merchants had significant 6.38 times higher knowledge than those of laborers (OR = 6.38, 95% CI = 1.64 – 24.74, *p* = 0.007) (Table 16). In terms of the biting time of *Aedes* mosquitoes, participants with the highest education at the secondary school had significantly 7.09 times higher knowledge than those with no education (OR = 7.09, 95% CI =1.14 – 43.96, *p* = 0.035); and merchants had significantly 4.44 times higher knowledge than those of laborers (OR = 4.44, 95% CI = 1.49 – 13.23, *p* = 0.007).

**Table 16.** Odds ratios of the knowledge on Zika with socio-demographic characteristics.

| Variable | Odd Ratios | 95%CI | P-value |
| --- | --- | --- | --- |
| **I. Zika is transmitted through mosquito bites** |  |  |  |
| **Age** |  |  |  |
| Age < 35 | Ref |  |  |
| Age 35-54 | 0.30 | 0.12 – 0.77 | 0.012* |
| Age >54 | 0.94 | 0.41 – 2.17 | 0.893 |
| **Occupation** |  |  |  |
| Laborer | Ref |  |  |
| Farmer | - | - | - |
| Merchant | 6.38 | 1.64 – 24.74 | 0.007* |
| Government/<br>State Enterprise officer | 3.37 | 0.89 – 12.72 | 0.073 |
| Company employer | 1.12 | 0.35 – 3.57 | 0.844 |
| Housekeeper | 3.89 | 0.37 – 41.33 | 0.260 |
| Others | 2.33 | 0.63 – 8.62 | 0.204 |
| **II. Biting time of *Aedes* mosquitoes** |  |  |  |
| **Education** |  |  |  |
| None | Ref |  |  |
| Primary school | 4.00 | 0.45 – 35.79 | 0.215 |
| Secondary school | 7.09 | 1.14 – 43.96 | 0.035* |
| High school | 3.13 | 0.51 – 19.09 | 0.217 |
| Bachelor degree | 3.33 | 0.53 – 21.03 | 0.200 |
| Higher than Bachelor degree | 2.70 | 0.45 – 16.22 | 0.278 |
| **Occupation** |  |  |  |
| Laborer | Ref |  |  |
| Farmer | - | - | - |
| Merchant | 4.44 | 1.49 – 13.23 | 0.007* |
| Government/<br>State Enterprise officer | 2.91 | 0.86 – 9.83 | 0.085 |
| Company employer | 1.16 | 0.43 – 3.12 | 0.773 |
| Housekeeper | 1.20 | 0.20 – 7.31 | 0.843 |
| Others | 3.26 | 0.98 – 10.88 | 0.055 |

\* Significant difference at $p < 0.05$

### Odds ratios of the knowledge on sterile mosquitoes with socio-demographic characteristics

For sterile mosquitoes, it was found that age, monthly income and house ownership had significant correlations with the familiarity of participants with the wording “sterile mosquitoes”. Participants aged more than 54 years had significantly 53% less familiar than those aged lower than 35 years (OR = 0.47, 95% CI = 0.28 – 0.78, *p* = 0.004) (Table 17). Participants with the income higher that 5,001 THB, were significantly two times up to almost five times more familiar with sterile mosquitoes than those with income less than 5,000 THB. Participants who were tenants had significantly two times higher familiarity than those of household owners (OR = 2.07, 95% CI = 1.30 – 3.30, *p* = 0.002). In term of the attitude towards sterile mosquitoes, participants with the incomes from 20,001 – 30,000 THB (US$ 578 – 867) had significantly 3.10 times better attitude than those with the incomes less than 5,000 THB (US$ 144) (OR = 3.10, 95% CI = 1.04 – 9.23, *p* = 0.042). When focused on overall rating scores, it was found that participants aged 35-54 years, rated 3.85 higher positive scores than those aged lower than 35 years (OR = 3.85, 95% CI = 1.27 – 11.71, *p* = 0.017). Participants living with more than two family members gave significantly lower scores than those living with less than two family members. When asked about willingness to purchase sterile mosquitoes with their own expenses, merchants had significantly 63% lower interest in purchasing sterile mosquitoes than those of laborers (OR = 0.37, 95% CI = 0.15 – 0.84, *p* = 0.019). Lastly, participants living with more than two family members had 1.85 times higher interest in purchasing sterile mosquitoes than those who lived with one to two family members (OR = 1.85, 95% CI = 1.02 – 3.33, *p* = 0.042).

**Table 17.** Odds ratios of the knowledge on sterile mosquitoes with socio-demographic characteristics.

| Variable | Odd Ratios | 95%CI | P-value |
| --- | --- | --- | --- |
| **I. Heard (familiar) about sterile mosquitoes** |  |  |  |
| **Age** |  |  |  |
| Age < 35 | Ref |  |  |
| Age 35-54 | 0.59 | 0.30 – 1.14 | 0.115 |
| Age >54 | 0.47 | 0.28 – 0.78 | 0.004* |
| **Monthly income (THB) (US \$1 = 34 THB)** | | | |
| < 5,000 | Ref |  |  |
| 5,001-10,000 | 4.75 | 1.57 – 14.37 | 0.006* |
| 10,001-15,000 | 5.14 | 2.00 – 13.24 | 0.001* |
| 15,001-20,000 | 2.77 | 1.03 – 7.45 | 0.043* |
| 20,001-30,000 | 3.54 | 1.28 – 9.74 | 0.015* |
| > 30,000 | 1.80 | 0.53 – 6.12 | 0.346 |
| **House ownership** |  |  |  |
| Owner | Ref |  |  |
| Tenant | 2.07 | 1.30 – 3.30 | 0.002* |
| **II. Good attitude toward sterile mosquitoes** |  |  |  |
| **Monthly income (THB) (US\$ 1 = 34 THB)** | | | |
| < 5,000 | Ref |  |  |
| 5,001-10,000 | 1.13 | 0.38 – 3.38 | 0.829 |
| 10,001-15,000 | 1.53 | 0.60 – 3.90 | 0.373 |
| 15,001-20,000 | 1.27 | 0.48 – 3.39 | 0.634 |
| 20,001-30,000 | 3.10 | 1.04 – 9.23 | 0.042* |
| > 30,000 | 0.92 | 0.29 – 2.91 | 0.891 |
| **III. Overall good rating scores for sterile mosquitoes** |  |  |  |
| **Age** |  |  |  |
| Age < 35 | Ref |  |  |
| Age 35-54 | 3.85 | 1.27- 11.71 | 0.017* |
| Age >54 | 1.79 | 0.63 – 5.08 | 0.273 |
| **Number of family members** |  |  |  |
| 1 - 2 | Ref |  |  |
| 3 – 4 | 0.11 | 0.01 – 0.90 | 0.040* |
| > 5 | 0.37 | 0.14 – 0.97 | 0.043* |
| **IV. Willing to pay for sterile mosquitoes at own costs** |  |  |  |
| **Occupation** |  |  |  |
| Laborer | Ref |  |  |
| Farmer | - | - | - |
| Merchant | 0.37 | 0.15 – 0.84 | 0.019* |
| Government/<br>State Enterprise officer | 0.58 | 0.23 – 1.48 | 0.257 |
| Company employer | 0.44 | 0.19 – 1.03 | 0.059 |
| Housekeeper | 0.88 | 0.23 – 3.34 | 0.845 |
| Others | 0.47 | 0.18 – 1.27 | 0.137 |
| **Number of family members** |  |  |  |
| 1 - 2 | Ref |  |  |
| 3 – 4 | 1.27 | 0.61 – 2.66 | 0.524 |
| > 5 | 1.85 | 1.02 – 3.33 | 0.042* |

\* Significant difference at $p < 0.05$

## Discussion

This study revealed that participants living in Bangkok were more familiar with dengue (85.25%) and this was much higher than those of chikungunya (39.75%) or Zika (33.75%). Our study was supported by Saminpanya and Jarujit who reported high level of knowledge of respondents towards dengue (71%) when compared to those of chikungunya (23%) [Saminpanya & Jarujit, 2022]. Dengue has been endemic for more than 60 years in Thailand [Nontapet et al., 2022], and it has been recognized as a major public health problem in Thailand [Thisyakorn et al., 2022] since the first major outbreak of dengue in Thailand in 1958 [Ooi & Gubler, 2009; Gubler, 1998]. Moreover, dengue virus infections usually occurred with the common severe presentation of hemorrhagic manifestations and occasional deaths from shock [Gubler, 1998], thus it remained the significant threat to the welfare of the Thai population [Khongwichit et al., 2018]. Although, chikungunya outbreak was also reported in 1958 [Khongwichit et al., 2021; Thaikruea et al., 1997] but little was known, so it received little attention as it had generally been regarded as self-limiting with few severe complications [Rianthavorn et al., 2010]. For Zika, the first report of the possible occurrence of Zika virus in Thailand was published in 1963 [Pond et al., 1963] and Zika virus has been circulating in Thailand since 2012 [Buathong et al., 2015]. However, since Zika virus infections represented a small proportion of ongoing flaviviral infections in Thailand [Khongwichit et al., 2018], with the exception of rare cases of more severe disease [Pinto-Díaz et al., 2017]. In addition, Zika was a relatively mild and self-limiting disease, which is often resolved without medical attention [Musso & Gubler, 2016], thus little was known for Zika, especially to general public.

In this study, participants had generally high knowledge on mosquito vectors and vector-borne diseases, i.e., they could identify at least one mosquito breeding site, they could correctly identify biting time of *Aedes* mosquitoes, and they considered dengue, chikungunya, and Zika as severe diseases although they did not experience virus infections by themselves. High level of knowledge has been related to the diffusion of information [Mejía et al., 2016] and media played a major role in the dissemination of public health knowledge [Boonchutima et al., 2017; Doblecki-Lewis et al., 2016; Arellano et al., 2015]. Television was the main source of information for participants in Thailand followed by health officers or village health volunteers. Al-Dubai et al. also showed that TV was the main source of information in Malaysia [Al-Dubai et al., 2013]; and Swaddiwudhipong et al. revealed an important role of health personnel in disseminating information and prevention methods of dengue in Thailand [Swaddiwudhipong et al., 1992]. Village health volunteers played an important role in facilitating effective health activities that increase awareness, motivation, involvement and monitoring health status within a community [WHO, 2020; Tejativaddhana et al., 2018; Kaewta et al., 2011]; and together with primary care units, they were the first point of contact with primary care and broader health system in Thailand [Nontapet et al., 2022].

In terms of prevention and control measures for vector control, covering water storage containers was the most common prevention measures responded by participants, followed by disposal of mosquito breeding sites such as discarded water containers or garbage, and weekly change of water in the storage containers. These results were supported by the study of Swaddiwudhipong et al. [Swaddiwudhipong et al., 1992]. High number of *Aedes* mosquitoes was presented in water storage, and large water storage represented 80% of *Aedes* breeding sites [Seng et al., 2008]. This could explain why people might be aware of clearing these containers [Kumaran et al., 2018]. However, when it comes to practices, although the majority of participants believed that covering water storage could help getting rid of mosquito breeding sites, but many of them never practiced it (47.75%). Kumaran et al. also found no correlation between knowledge and practices [Kumaran et al., 2018]. However, our results were contradicted with Al-Dubai et al. who found an association between knowledge and practices in dengue control [Al-Dubai et al., 2013]. When focus on attitudes, although the majority of participants had good attitudes in many aspects, such as regularly change water in the water storage containers, sleep under the bed net, prevent themselves from mosquito bites; but when it comes to the elimination of mosquito breeding sites, many of them still believed that it was a sole responsibility of the health officials. This finding was supported by Kamaran et al. who showed that people were less likely to take vector control measures despite of its benefits when government had performed vector control measures for a long time and people believed that it was the government’s responsibility [Kumaran et al., 2018].

When focused on knowledge of sterile mosquitoes, many participants knew nothing about sterile mosquitoes (45.50%) but some of them had some knowledge on sterile mosquitoes (37.75%), such as how to sterilize mosquitoes, benefits of sterile mosquitoes, difference between sterile and wild mosquitoes, etc. Our study indicated that municipal officials were the major source of information about sterile mosquitoes followed by leaflets or flyers, and village broadcast towers. More efforts have to be done in order to deliver more information and knowledge on sterile mosquitoes through previously mentioned channels of communication in order to reach more people in the community prior to the release or implementation of sterile mosquitoes in the fields. Moreover, various media channels, i.e., TV and online media, were needed in order to reach more people and general public. Good communication strategy is essential in SIT field trials for soliciting acceptance of the community [Stefopoulou et al., 2021; Genovesi et al., 2011]. When focused on attitudes towards sterile mosquitoes or an application of sterile mosquitoes to reduce the mosquito vectors of dengue, chikungunya and Zika, although the majority of people showed positive attitudes towards sterile mosquitoes (47%), i.e., they were interested in having the sterile mosquitoes released in their home or community, and they believed that it could reduce chemical use and reduce the *Aedes* mosquito populations in their home or community, but a non-negligible percentage of people was still uncertain about sterile mosquitoes (33%). When asked about the willingness to pay for sterile mosquitoes, the majority of participants did not want to purchase the sterile mosquitoes by their own expenses as they preferred to receive them free of charge from the government. For those few of them who were willing to pay, only 9.0-9.5% of them could afford to purchase the sterile mosquitoes with the price of 1-2 THB (US$ 0.03-006) per sterile mosquito.

When asked about health seeking behavior, when participants of their family members got bitten by mosquitoes or got sick, the majority of them primarily preferred self-medication at home or preferred buying medicines for their own use (49-55%) before going to see a medical doctor (18.75%) or visiting a specialist (10.12%). Their major expense was about 100 THB (US$ 2.86) for medicine or about 350 THB (US$ 10) for the cost of medical diagnosis. Kumaran et al. reported that self-medication was mostly chosen by people in Cambodia when they were sick; and Piroonamornpun et al. reported 95.2% of patients in Thailand had self-medication with paracetamol and antibiotics prior to seeking other health care due to an easy access to over-the-counter drugs [Kunaran et al., 2018; Piroonamornpun et al., 2022]. When focused on other expenses related to vector control, it could be ranged from 100 THB (US$ 2.86) for mosquito repellent spray or up to 2,500 -5,000 THB (US$ 71.50 – 5,000) for mosquito traps. These expenses caused economic burden to people since the minimum daily wage in Thailand ranged from 328-354 THB (US$ 9.12-9.84) [ASEAN Briefing, 2023]; thus the cost for medical treatment of 350 THB was almost already surpassed the daily minimum wage. As many participants were laborers whose incomes largely depended on their presence at work, therefore when they were absence from work due to illness or medical visits, it could have an impact on their quality of life or their family members in the case that they were head of the family.

When asked about an impact of the release of sterile mosquitoes in terms of the environment, economic, social and quality of life, the majority of participants showed positive perception and believed that they could benefit from it. However, when asked about an impact on the reduction of mosquito population or chemical uses for vector control, good relationship with neighbors, satisfaction of living condition, and awareness of information from release of sterile mosquitoes, some level of certainly was observed among participants. This could be explain by the fact that the release of sterile mosquitoes had not yet been implemented in the fields so that people could hardly imagine or think about any impacts that could happen to them or to their communities.

## Data Availability

The data set will be submitted to public data base website before publication.

## Acknowledgement

The authors would like to thank the staff of the Department of Health, Bangkok Metropolitan Administration (BMA) for their field accompany and their assistance with the questionnaire management.

